# DeepOS: pan-cancer prognosis estimation from RNA-sequencing data

**DOI:** 10.1101/2021.07.10.21260300

**Authors:** M. Pavageau, L. Rebaud, D. Morel, S. Christodoulidis, E. Deutsch, C. Massard, H. Vanacker, L. Verlingue

**Affiliations:** Drug Development Department (DITEP), Gustave Roussy - Cancer Campus, Villejuif, France; CentraleSupelec, Gif sur Yvette, Paris Saclay, France; INSERM UMR1030, Molecular Radiotherapy and Therapeutic Innovations, Gustave Roussy - Cancer Campus, Villejuif, France; UPS, University Paris Saclay; Centre Léon Bérard, Lyon, France

**Keywords:** neoplasm, prognosis, deep learning, transfer learning, RNA-sequencing

## Abstract

RNA-sequencing (RNA-seq) analysis offers a tumor-centered approach of growing interest for personalizing cancer care. However, existing methods – including deep learning models – struggle to reach satisfying performances on survival prediction based upon pan-cancer RNA-seq data. Here, we present DeepOS, a novel deep learning model that predicts overall survival (OS) from pan-cancer RNA-seq with a concordance-index of 0.715 and a survival AUC of 0.752 across 33 TCGA tumor types whilst tested on an unseen test cohort. DeepOS notably uses (i) prior biological knowledge to condense inputs dimensionality, (ii) transfer learning to enlarge its training capacity through pre-training on organ prediction, and (iii) mean squared error adapted to survival loss function; all of which contributed to improve the model performances. Interpretation showed that DeepOS learned biologically-relevant prognosis biomarkers. Altogether, DeepOS achieved unprecedented and consistent performances on pan-cancer prognosis estimation from individual RNA-seq data.

## Introduction

Among patients diagnosed with cancer, prognosis estimation is often required to draw a risk profile and adapt treatment accordingly. Currently recommended prognostic and predictive biomarkers that drive cancer care management usually combine several items, such as: individual characteristics (e.g. age, gender, ECOG status), tumor characteristics (e.g. tumor stage, localization and number of metastasis), serum markers (e.g. albumin, LDH, CRP) and eventually tumor molecular features (e.g. PD-L1 expression, *BRCA* loss-of-function, *ERBB2, EGFR, BRAF, ALK* mutations, *NTRK* fusion) ^1–6^. Such scoring systems mainly stratify patients into low- or high-risk groups, defining therapeutic procedures to be followed. More recently, the growing interest in high-dimensional multi-omics data in assisting clinicians on treatment decision has brought forward the high potential of tumor RNA-sequencing (RNA-seq) on studying the link between tumor gene expression and patient survival outcome in a personalized way ^7^. RNA-seq provides gene expression quantifications of the whole transcriptome (transcripts of more than 20,000 protein-encoding genes) or of preselected transcripts of interest (targeted sequencing), bearing the underlying hypothesis that each tumor gene expression profile mirrors the tumor aggressiveness and potential behavior in response to a particular treatment and therefore, should correlate with overall survival (OS).

Since each tumor is unique in its complexity, an almost-infinite number of gene expression combinations could be expected; which drastically overcomplicates any prediction task based on RNA-seq data. Several teams recently intended to predict individual OS from RNA-seq analyses of multiple cancer types obtained from the Cancer Genome Atlas (TCGA) dataset ^8^, using machine and deep learning ^9–14^ (summarized in Supplementary Table 1). Model architectures included Random Forest, Cox regression with Lasso penalization, Multilayer Perceptron (MLP), Convolutional Neural Networks, Auto-Encoder with Cox loss function, among others. Those models prediction performances in validation or test cohorts were often limited, close to 0.60, and rarely exceeded 0.62 of median concordance-index (C-index) on pan-cancer predictions. C-index is a popular metric that evaluates the ability of a model to rank survival predictions within a particular cohort rather than the difference between the predicted and the observed values (with C-indexes of 0.50 and 1.00 respectively corresponding to random and perfect order of predictions) ^15,16^. Yet, prognosis estimation from pan-cancer RNA-seq data should be feasible since each tissue and each tumor type express their own transcriptomic signatures.

With machine and deep learning, over-fitting can arise when the training dataset has greater number of dimensions (variables) than number of samples available (the size of the training set). Over-fitted models generally fail to generalize decently ^17^. In the case of RNA-seq, the large dimensionality (e.g. > 20,000 gene expressions) requires massive amounts of training data, which is tricky to obtain when applied to cancer and which presumably lacked to the above-mentioned models (total number of samples comprised between 953 and 11,854). We hypothesized that OS prediction using supervised deep learning on pan-cancer RNA-seq data as inputs would benefit from (i) starting with reducing input dimensions using prior biological knowledge and (ii) increasing the size of the training set, using a transfer learning strategy. Transfer learning consists in pre-training a model on a task that is related, but not strictly identical to the final question, and for which a larger number of samples is available ^18^. We therefore (i) filtered whole transcriptome expressions to reduce inputs dimensionality and (ii) designed an MLP neural network that we pre-trained to predict the organ of origin from large healthy and cancer RNA-seq data and fine-tuned to predict OS from pan-cancer RNA-seq data (Figure 1). We additionally characterized the parameters that influenced the model performances, including the gene list selection, the size of the training set, the type of survival loss function for training, the duration of OS, the cancer type and the genes the most implicated into the model prediction.

**Figure 1:**
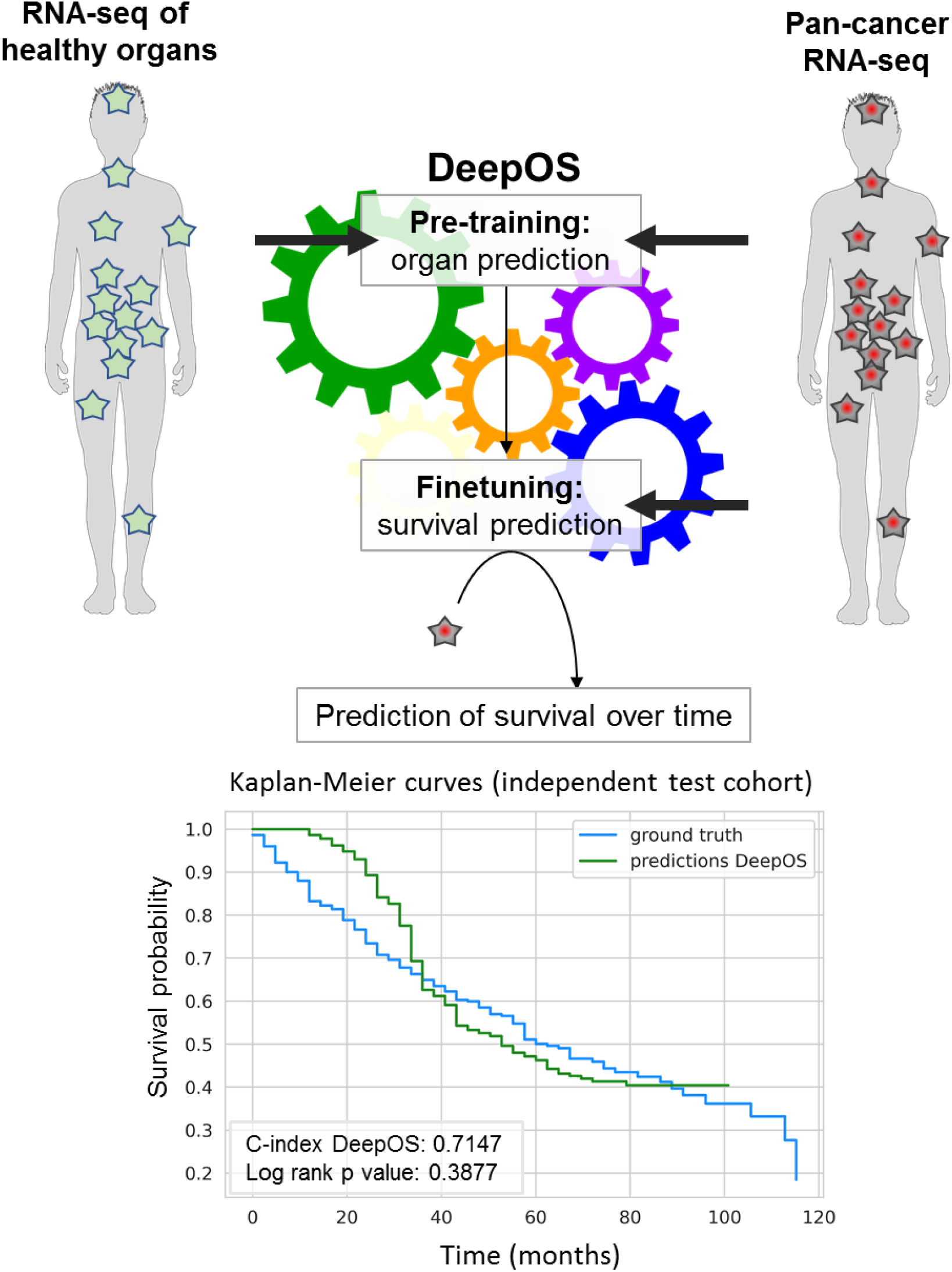
Graphical abstract: Pipeline description of DeepOS. First, the model is pretrained to predict the organ of origin of both healthy and tumor tissues. Then, the model is fine-tuned on survival of the pan-cancer RNA-seq cohort. DeepOS is a multilayer perceptron neural network model, that uses the RNA-seq expression of 4,499 cancer and immune genes as inputs. DeepOS outputs a probability of survival per time intervals (in the example, one interval represents 72 days). This allows training DeepOS on censored survival data. Survival is estimated by the first interval meeting the probability = 0.5. On the example, a 50% risk of death is predicted to occur at the 14^th^ interval, which corresponds to 33 months.

## Results

### Gene selection

We first intended to reduce the dimensions of our input dataset by selecting genes of interest, which are known to be implicated in cancer initiation, progression, dissemination or response to treatment. We merged gene lists obtained from the Molecular Signatures Database (MSigDB, relative to hallmarks of cancer) ^19^, and the LM22 immune gene signatures ^20^. After removal of duplicates and genes associated with no expression values within our dataset, we obtained 4,499 genes (Supplementary Table 2).

### Pilot overall survival prediction task: without pre-training

We designed a pilot experiment of survival estimation starting with only tumor RNA-seq data. We retrieved all The Cancer Genome Atlas (TCGA) pan-cancer RNA-seq raw data publicly-available on February, 11, 2019, via recount2 ^21^ and selected samples associated with annotated survival outcomes (Figure 2a). We excluded uninformative patients who were censored during the first half of the total duration of the follow-up and the top 5% of patients with the longest OS, considering them cured by surgery. Altogether, we collected 6,529 RNA-seq samples from 33 tumor types fulfilling the criteria, among which 54.8% were censored during second half of the follow-up.

**Figure 2:**
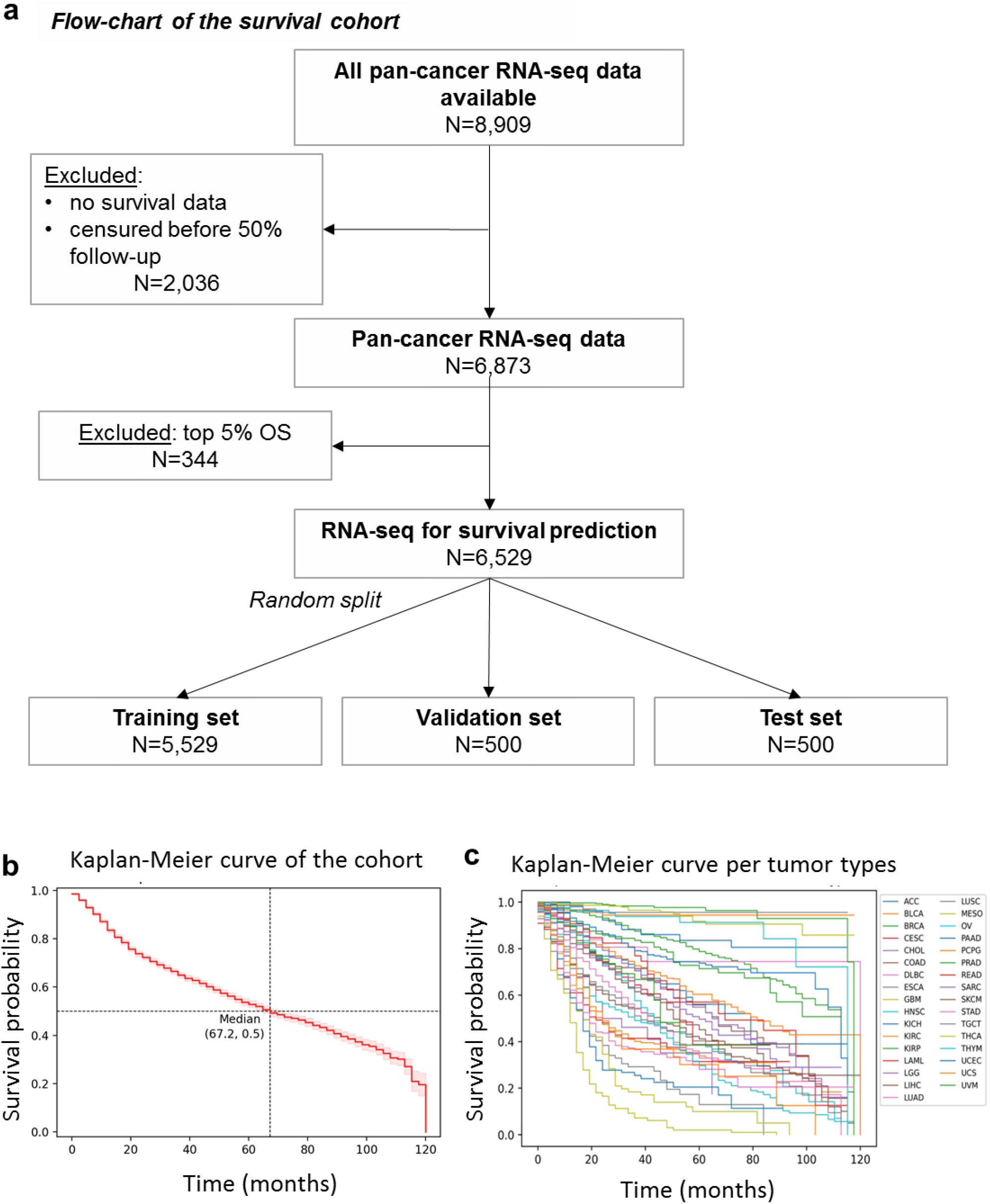
Pan-cancer survival data description. **a**, Flow-chart of the survival cohort. Pan-cancer RNA-seq and survival and clinical data were retrieved from the TCGA dataset. After selection of the 6,529 samples fulfilling the selection criteria, we used an 80%/10%/10% random split rule to create the training, validation and testing datasets. **b**,**c**, Kaplan-Meier survival curves of the whole cohort (**b**) and per tumor types (**c**). TCGA study abbreviations, median overall survival per tumor type and 95% confidence intervals are detailed in Supplementary Table 3.

Based on this dataset, the pan-cancer median OS was 67.2 months (95% confidence interval 95%CI [64.8;72.0]) (Figure 2b), and highly depended on the tumor type (Figure 2c). Glioblastoma, esophageal cancer, mesothelioma and pancreatic cancers were associated with the worst prognoses (median OS of 12.0 months for glioblastoma and 16.8 months for the three others); on the other hand, median OS was not reached after a 120-months follow-up for five tumor types (chromophobe and papillary renal carcinoma, pheochromocytoma, testicular cancer and thyroid cancer) (Supplementary Table 3).

We randomly assigned each sample to either a training set (N=5,529), a validation set (N=500) or a test set for final evaluation (N=500). Splitting was well-balanced considering the fraction of censored patients (0.546, 0.536 and 0.546, respectively within training, validation and test cohorts), median OS (67.2 months, 72.0 months and 62.4 months, respectively) and diversity of cancer types (Supplementary Table 4 and Supplementary Figure 1).

To train our models, we transformed survival data into survival probabilities per time interval and thus, could implement the classical mean squared error (MSE) loss function. Survival probabilities were set to 1 for intervals during which a patient is alive, and to 0 when a patient is deceased. Censored intervals were ignored to calculate the loss. Ignoring censored intervals in MSE allowed the model to be trained only on observed time intervals for each patient. The model learned a probability of survival for each patient individually, using classical methods in deep learning for multiclass classification. To decipher whether this approach could be competitive, we compared the performances of DeepOS to DeepSurv, a state-of-the-art deep learning model based on Cox-loss to train on survival data ^22^.

We used a Tree-structured Parzen Estimator (TPE) ^23^ algorithm to explore hyper-parameters and to select the best model upon the highest C-index obtained on the validation set. After training on 5,529 pan-cancer RNA-seq samples, the best model reached a C-index of 0,744 on the validation set, with cross-validation C-index mean of 0.63 and standard deviation of 0.09. The best model had five hidden layers, each of them having a dropout of 0.015, and L1 and L2 penalization of 0.007 and 0.0009, and trained with a learning rate of 0.00003.

On a final and previously unseen test cohort, this model achieved a C-index of 0.707 on predicting patient survival from their pan-cancer RNA-seq data. This model surpassed DeepSurv performances on the same split data (C-index of 0.606 on validation and test sets).

### Learning curves of survival prediction

To study how the number of samples within the training set influenced the model performances, we repeated the survival training task with escalating number of samples composing the training cohort, without modifying the validation set.

Learning curves indicated that C-indexes reach a steady state for training cohorts containing at least 2,000 RNA-seq samples (Figure 3a). The best training set C-index (0.80) was achieved with 1,000 samples; although the difference between training- and validation-related C-indexes indicated that the model was subject to overfitting (Figure 3b). Overfitting defines a model that learns too perfectly from a training set so that it fails to generalize adequately on unseen additional data. According to our results and consistently with what was previously described ^24,25^, overfitting tends to be reduced by increasing the number of samples within the training set (Figure 3b). We therefore hypothesized that a transfer learning strategy could benefit our model since it could indirectly expand the training dataset through pre-training on a similar task.

**Figure 3:**
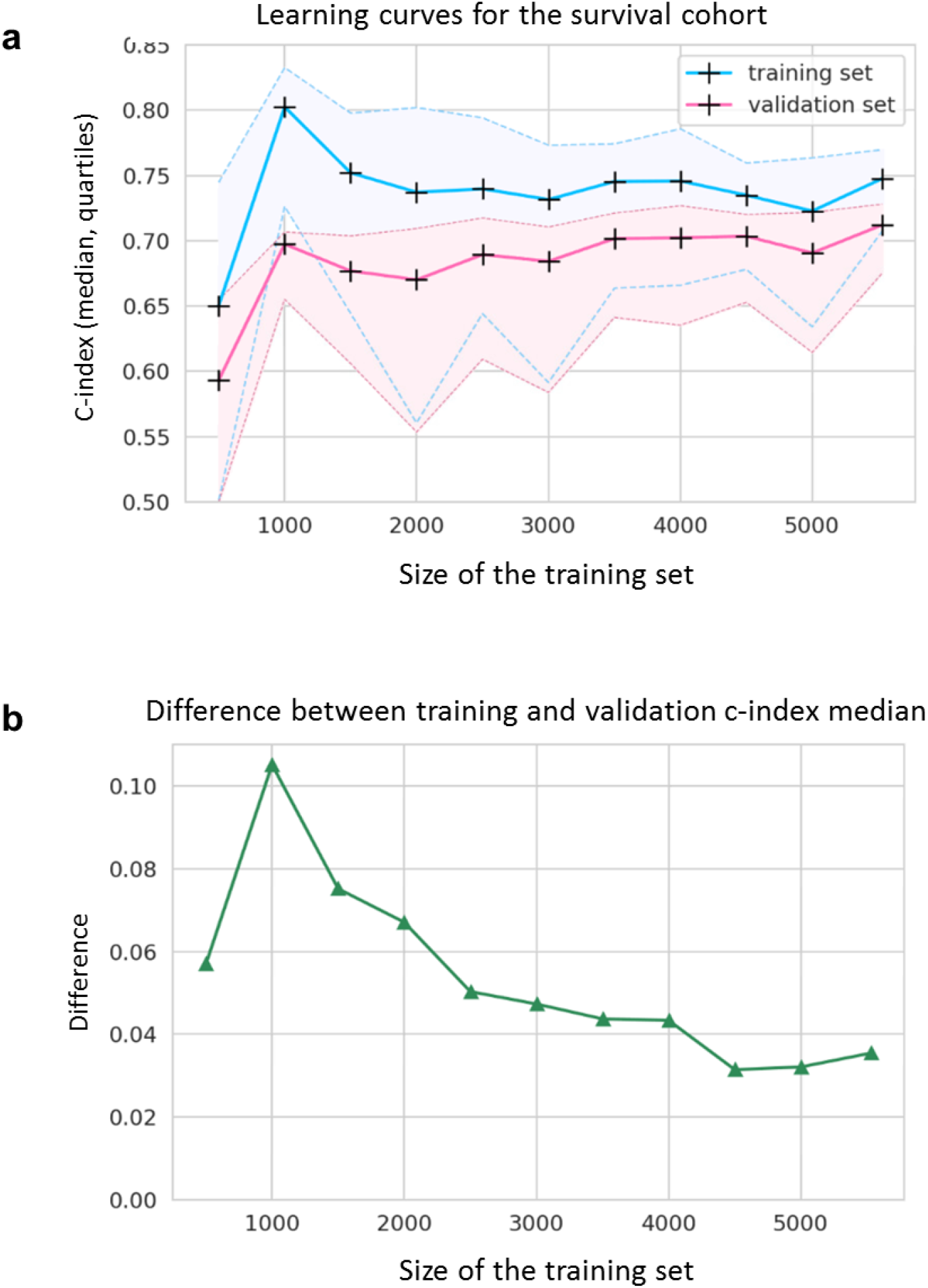
Survival learning curves. a,b, Learning curves for the survival cohort represented by line charts of the median, 1^st^ and 3^rd^ quartiles of the C-indexes on the training (blue) and the validation (pink) datasets (a) and the resulting difference between training and validation median C-indexes (b) according to the size on the training set (from 500 to 5,529 samples, with steps of 500 samples). C-index = concordance index.

### Transfer learning: data collection for the pre-training task

Since OS was highly related to tumor type (Figure 2b), we assumed that learning to predict the organ of origin from a larger cohort could improve the overall estimation of survival duration. We therefore chose to pre-train our model on the prediction of the organ of origin from the RNA-seq expression data of the 4,499 selected genes using either healthy or tumor tissue (Figure 4a). Healthy organs data were obtained through recount2 from the Genotype-Tissue Expression (GTEx) project ^26^. We additionally retrieved all the TCGA gene expression data of tumor samples (including those not associated with survival data). Each tumor type was aligned with its organ of origin (for example, kidney chromophobe, clear cell carcinoma and papillary cell carcinoma were all considered as kidney tissue).

**Figure 4:**
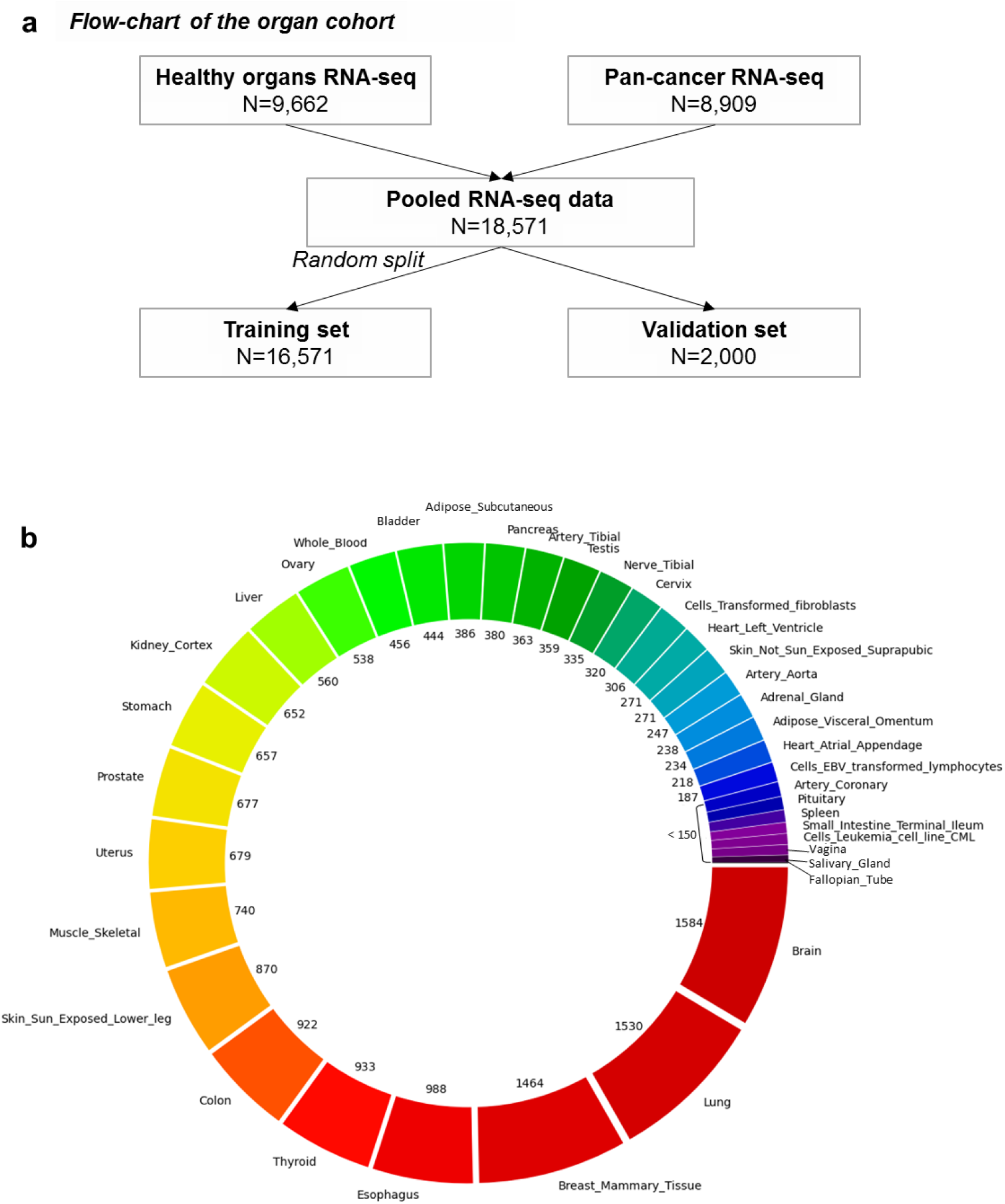
The organ cohort. a, Flow-chart of the organ cohort used to pre-train DeepOS. Healthy organs RNA-seq were obtained from the GTEx project, while pan-cancer RNA-seq data were obtained from the TCGA dataset. Samples were randomly assigned to either the training or the validation cohort with a 90%/10% split. b, Distribution of organ types across pooled RNA-seq data used for pre-training. The number alongside each feature refers to the number of patients.

Altogether, we collected 18,571 RNA-seq samples from 38 distinct human tissues (Figure 4b). The most represented organs were brain (8.5% of samples), lung (8.2%) and breast tissue (7.9%). We randomly divided the samples into two distinct sets for training (16,571) and validation (2,000). Splitting was well balanced and both sets harbored samples belonging to the 38 types of tissue.

### Pre-training on organ prediction

We pre-trained the first section DeepOS, considered as “low abstraction”, on organ prediction. DeepOS is an MLP neural network, that takes gene expression values in transcripts per million (TPM) as inputs, and outputs either organ classification or survival probabilities (Supplementary Figure 2). DeepOS architecture comprises hidden layers composed of stacked units of dense layers, Rectified Linear Unit (ReLU) activations, dropout effect penalization, L1 and L2 regularization, and batch normalization.

All the models tested reached very high validation performances to predict the organ of origin (mean hyper-parameter search accuracy = 0.849 and standard deviation = 0.292), with best model reaching an accuracy of 0.9835, precision of 0.9842, recall of 0.9835 and F1-score of 0.9836 (Supplementary Figure 3). The best organ-specific model had four layers, with a dropout rate of 0.080, a L1 and L2 regularization parameters respectively of 0.0013 and 0.0037 and was trained with a learning rate of 0,8462. We did not perform a test set evaluation as this step was only used to select the best pre-trained model to fine-tune.

### Fine-tuning on survival prediction

To implement our transfer learning strategy, we then fine-tuned DeepOS on survival prediction based on the pan-cancer cohort described above. The low abstraction section of DeepOS was frozen during the first fine-tuning step and unfrozen during the second fine-tuning step (Supplementary Figure 2). Similarly to the pilot task performed without pre-training, we selected the best version of the fully trained (pre-training and fine-tuning) model based on the highest C-index obtained on the validation set, which reached 0.738 (cross-validation C-index mean = 0.688, standard deviation = 0.058).

On the unseen test cohort, this DeepOS model achieved a C-index of 0.715 and an area under the survival ROC curve (AUC) of 0.752 (Figure 5a and 5b). Transfer learning improved survival prediction of +0.9% and +0.4% respectively for DeepOS C-index and mean AUC, which are noteworthy upgrades for models with performances >0.70 on this type of task. In addition to the organ-specific features, the best model architecture had six layers, with a dropout rate of 0.027 and L1 and L2 regulation parameters respectively of 0.0025 and 0.0069.

**Figure 5:**
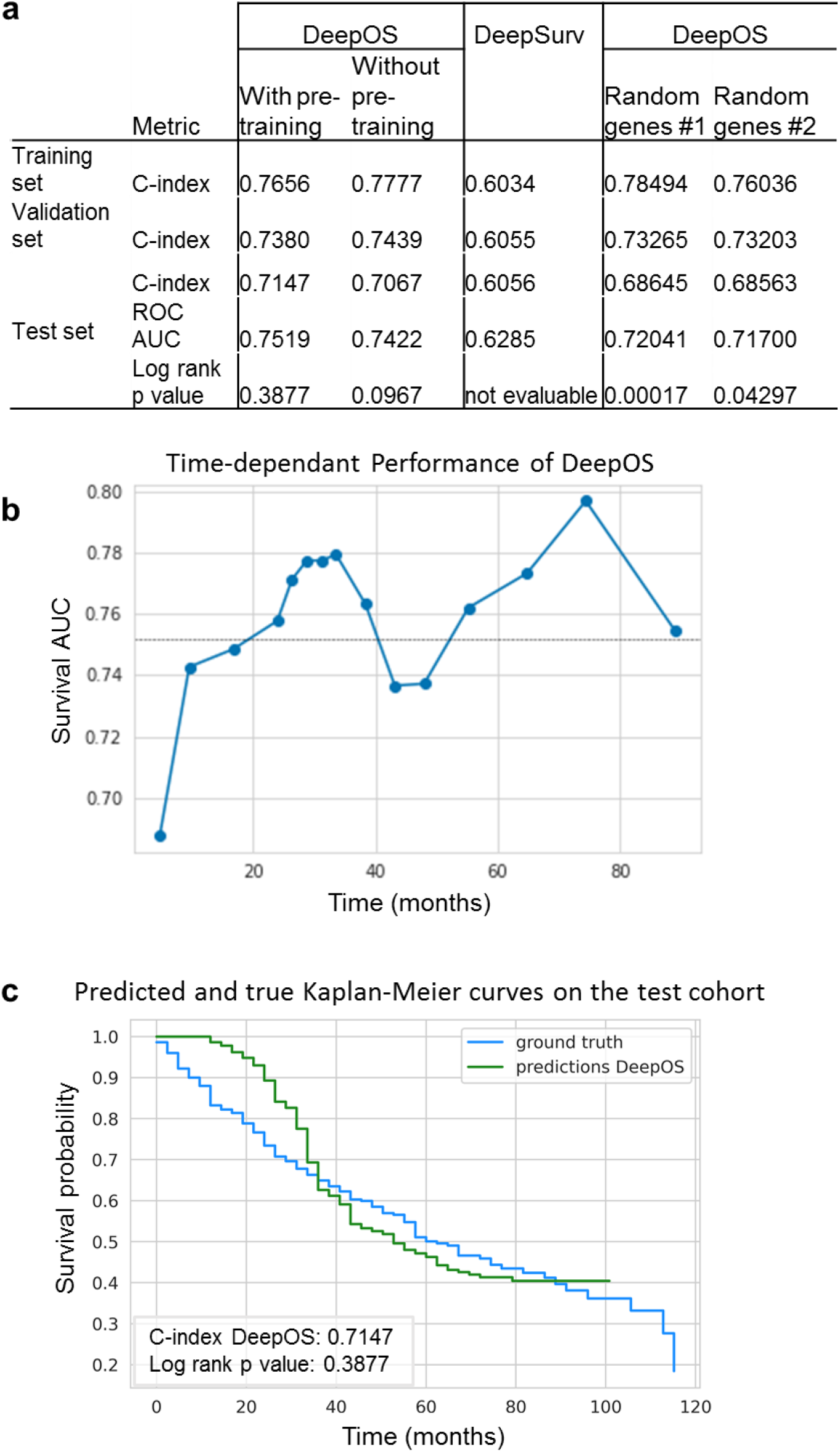
DeepOS results. a, Summary of the performances obtained with DeepOS with and without pre-training on organ prediction, DeepSurv and DeepOS with pre-training while using random gene selection. Performances on the training, validation and unseen test set are depicted, based on the same data split. The gene set lists used for DeepOS predictions are detailed in Supplementary Table 2. b, Line chart of the survival Area Under the ROC Curve (AUC) according to time for DeepOS predictions on the test set. The grey vertical line refers to the mean of all AUC = 0.752. c, Kaplan-Meier survival curves of OS probability over time, either predicted from DeepOS (green) or observed (blue) within the test cohort. The predicted curve stops shortly after 100 months of OS, which corresponds to the longest OS prediction by DeepOS when analyzing the test cohort. Log-rank p-value = 0.39 indicates the absence of statistical difference between the two Kaplan-Meier curves. C-index: Concordance index; OS: overall survival.

### DeepOS according to patient OS

To study whether DeepOS can be used to predict pan-tumor survival, we generated a predicted Kaplan-Meier survival curve and compared it to the true survival curve of the test cohort (Figure 5c). We noticed that there was no significant difference between DeepOS prediction over time and the ground truth (log-rank p-value: 0.388). When compared to predictions without pre-training, transfer learning improved the accuracy of the model over time (Supplementary Figure 3a, log-rank p-value: 0.097 on the test set without pre-training). When applied to the training and validation cohorts, we similarly observed apparent closeness between predicted and observed curves for OS comprised between 30 and 100 months (Supplementary Figure 4b,c), although we repeatedly noticed divergences of the curve slopes for shorter OS, with DeepOS behaving over-optimistic as compared to reality.

To further evaluate DeepOS performances according to survival duration, we generated ten subgroups of 50 patients ranked by OS on the test cohort, and computed the C-indexes of each subgroup. We could indeed observe that DeepOS performed modestly for survival predictions of patients deceased between 1 and 20 months (C-index <0.60; <0.55 without pre-training) (Supplementary Figure 5a). This observation was not associated with underrepresentation of such population within the training set (Supplementary Figure 5b).

### DeepOS according to tumor type

Among the 29 cancer types that contained at least 3 uncensored samples, 26 (89.7%) displayed a C-index >0.50 which corresponds to better than random prediction and 12 (41.4%) had a C-index >0.72 on the test set, including two that reached the perfect score of 1 (adrenocortical carcinoma and uveal melanoma), although these were composed of only 5 and 4 patients within the test set, respectively (Figure 6a). Despite our previous observation, DeepOS was able to perform reasonably well on four out the five tumor types displaying a median OS <20 months, with C-indexes comprised between 0.55 and 0.74 (Supplementary Table 5). Transfer learning unchanged or improved survival predictions for 20 (69.0%) tumor types, up to +27,3%, as assessed by C-index calculation (Supplementary Figure 6, Supplementary Table 5).

**Figure 6:**
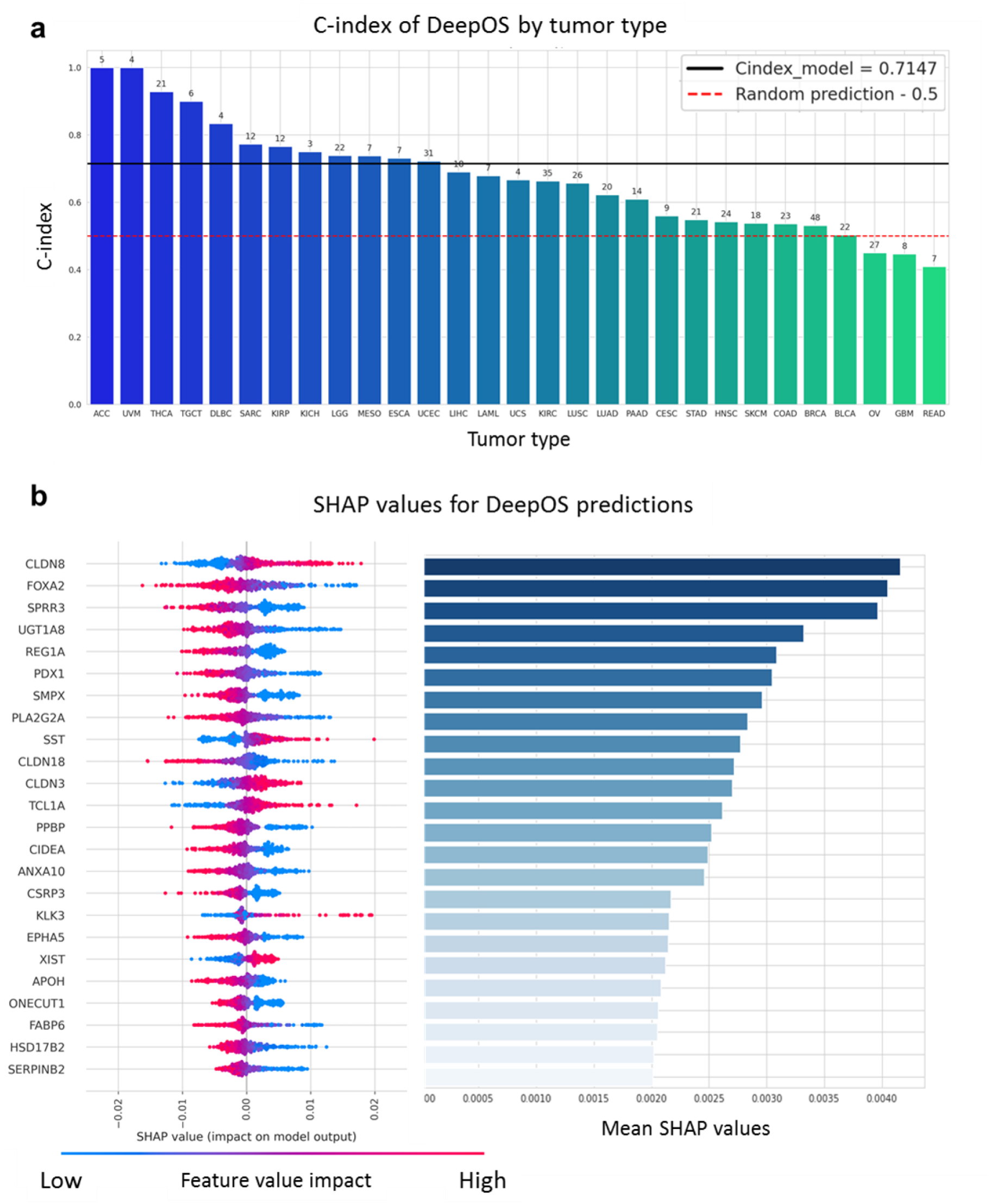
DeepOS interpretation. a, Bar chart of the C-indexes of DeepOS according to the tumor types. We considered tumor types within the test cohort with at least three uncensored samples. The red dotted line indicates a C-index of 0.50 (random prediction). The black line indicates a C-index of 0.715, which refers to the median C-index of DeepOS pan-cancer predictions. Patient numbers for each cohort are represented above the bars. b, The importance of DeepOS input gene is represented by a mirror bar chart of the SHapley Additive exPlanations (SHAP) values. SHAP values for individual predictions are plotted on the left panel. Genes are ranked by mean SHAP values as reported on the right panel. A high positive feature value (pink to the right) means that an increased expression of the gene is related to a reduced OS prediction, whereas a low positive feature value (blue to the right) means that a decreased gene expression is liked to a reduced OS prediction. Each dot represents an individual prediction.

### DeepOS according to gene selection

We then evaluated if the gene set selected on prior biological knowledge helped DeepOS performance. We thus trained models similar to DeepOS using two distinct and random selections of 4,499 genes (excluding the ones used in DeepOS) (Supplementary Table 2). Using random genes as inputs impaired the model performances, achieving C-index of 0.686 on the test set for both selections #1 and #2 (Figure 5a). Predicted Kaplan-Meier survival curves were significantly different to the ground truth (log-rank p-value: 0.00017 and 0.043 respectively for random selection #1 and #2) (Supplementary Figure 7). Gene selection based on prior-knowledge thus contributed significantly to DeepOS generalization performance.

### DeepOS according to gene expressions

To finally better characterize which genes were the most important for DeepOS predictions, we estimated the mean SHapley Additive exPlanations (SHAP) value for each gene. SHAP provides an interpretation of the importance attributed by the algorithm to each feature ^27^.

Among the 4,499 genes, 106 had a mean absolute SHAP value >0.001 and 24 genes had a mean absolute SHAP value >0.002, which we considered as the most important for DeepOS predictions (Figure 6b, Supplementary Figure 6b). We compared the effect of gene expressions importance and direction for the model decision with known biological findings. The most important gene was *CLDN8*, encoding for the Claudin-8 protein, with low expressions related by the model to poor OS. Claudin-8 is a transmembrane protein that constitutes tight junctions between epithelial cells; conversely, its downregulation has been previously related to tumorigenesis and epithelial-mesenchymal transition, and has been proposed as a biomarker of bad prognosis in several tumor types ^28–31^. Gene expression of *FOX2A* was the second most important feature, with high expression values correlated with poor OS. This is also consistent with the known role of FOX2A, a transcription factor promoting proliferation and epithelial-mesenchymal transition in multiple cancer types ^32–34^. Small proline-rich repeat protein 3 (*SPRR3*) ranked third among the most important genes for DeepOS, with high expressions related to poor OS prediction. Again, independent studies confirmed that high SPRR3 tumor expression was associated with significantly decreased survival, notably in pancreatic cancer, and with resistance to radiation therapy in head and neck cancer ^35,36^. In addition, poor OS were mainly associated by DeepOS to high expressions of *UGT1A8, REG1A, PDX1, SMPX, PLA2G2A, CLDN19, PPBP, CIDEA, ANXA10, CSRP3, EPHA5, APOH, ONECUT1, FABP6, HSD17B2* and *SERPINB2* and to low expressions of *SST, CLDN3, TCL1A, KLK3* and *XIST*, although the orientation (positive or negative correlation) could vary across tumor types (Supplementary Figure 8).

## Conclusions, discussion

We have developed a deep learning model, DeepOS, to estimate OS duration from pan-cancer RNA-seq data based on a transfer learning strategy that allowed us to enlarge our pre-training dataset with healthy tissue samples. Transfer learning improved prediction performances by limiting overfitting. The pre-training task consisted in predicting the organ of origin with very high accuracy, precision and recall performances (0.984 each on the validation set). For survival prediction, DeepOS reached a median pan-cancer C-index of 0.72 on an independent and previously unseen test set and a mean survival AUC of 0.76. DeepOS can output a discrete estimation of individual survival, which enables to plot Kaplan-Meier survival curves from individual predictions. Doing so, we could confirm that DeepOS survival predictions over time were not different from the ground truth in the test set (log-rank p-value 0.388). We also validated that the pre-training step increased the performance of the model.

We also found a way to use the mean square error, a classical loss function in deep learning, to train DeepOS on survival data. We used DeepSurv as an internal comparator using the same training, validation and testing data split. DeepSurv is a deep neural network trained with a Cox proportional hazards loss function, which is considered a state-of-the-art method for survival prediction ^22^. DeepOS significantly outperformed DeepSurv model performances (DeepSurv C-indexes were comprised between 0.60 and 0.61, and AUC of 0.63 on the test set, similarly to other published models for such task; Supplementary Table 1).

Our model has limitations. Firstly, as compared to the survival prediction task ran without pre-training, one could argue that the transfer learning strategy only slightly improved the model performances (+0.9% on the test C-index). However, the gain was robustly observed across all the metrics computed on the test dataset (C-index, mean AUC, log-rank p-value), which suggests that the model indeed benefited from the pre-training step. Besides, even minor upgrading in performance is challenging to obtain for C-index values above 0.70 for such task. Our study nevertheless supports a benefice of using large tumor RNA-seq datasets with survival observation.

Secondly, we noted over-optimism in DeepOS predictions for short survival durations (mainly <20 months). However, it did not negatively impact the model performances within tumor types of unfavorable prognosis. For example, the C-index within the mesothelioma and the esophageal carcinoma cohorts reached 0.74, while median OS was below 20 months (16.8 months each). A possible explanation is that patients with short survival have poor prognosis factors such as tumor location, poor general condition or comorbidities that are missed by using only RNA-seq data from a microscopic tumor sample. Analyzing the influence of time on models performance should be generalized to better define the application framework of deep learning models for such task.

Finally, the pan-cancer TCGA RNA-seq dataset that we used for our study was built mostly upon primary tumor samples obtained from surgical resection of localized or locally-advanced diseases. This is highlighted by the observed median OS for several cancer types longer than expected for metastatic stages at diagnosis. It is nevertheless possible that DeepOS learned from the metastatic potential of the tumor samples. This is supported by the detail of the genes with the highest importance for the algorithm, which are mainly related to cancer progression and epithelial-mesenchymal transition and thus, to cancer dissemination. However, further refinement and validation studies are warranted to statue on the generalizability of the model in metastatic cancers.

To our knowledge, DeepOS is the best performing model on individual pan-cancer survival prediction based on gene expression alone (Supplementary Table 1). Other approaches have proposed clustering analysis from RNA-seq to identify groups of patient with similar prognosis ^12 37^. *Thorsson et al*. could identify six immune subtype features from TCGA pan-cancer data comprising RNA-seq, miRNA-seq and exome sequencing data ^37^. They rigorously characterized immune subtypes associated with good and poor prognosis, although pan-cancer performances were modest with a median pan-cancer C-index visually lower than 0.60. In addition to transfer learning, DeepOS comprised methodological adaptations that we believe have permitted this upgrading. First, we transformed survival data into time interval survival probabilities, so that censored time intervals did not influence the loss function calculation. Thus, we could train on the mean squared error. We also reduced input dimensions by applying prior knowledge on the biology of cancer and immunity to limit overfitting due to irrelevant genes for our task, which contributed to improve the model predictions.

Overall, our study demonstrated and/or validated that (i) predicting survival outcomes from pan-cancer RNA-seq data is feasible and can achieve decent performances, (ii) transfer learning can reduce overfitting, and (iii) partially censored survival data can be used to train supervised deep learning models with standard loss functions. DeepOS offers a promising proof-of-concept that prognosis estimation among patients affected with various types of cancer can be personalized beyond classical score calculations. It provides a more tumor-centered way to estimate the disease aggressiveness and perhaps, to estimate its sensitivity to multiple therapeutic options.

## Methods

### Objectives

This study aimed at predicting the survival of patients affected by various tumor types from their gene expression analysis. This is a classical task with gold-standard datasets that we used to evaluate methodological improvements (Supplementary Table 1). We have developed a new format of survival data to train deep learning models, a prior-knowledge based dimension reduction and a transfer learning strategy. We hypothesized that these methods should help model performance and interpretability.

### Labels – Survival

We used the publicly available survival data of the TCGA database from *Liu et al*. ^38^. The top 5% of patients with the highest overall survival were removed because they were considered cured (by surgery, as their overall survival was higher than nine years). Patients with no follow-up were also removed (i.e. 0 days or survival status not known). Early censored patients had poor relevance for the training; we thus removed patients censored before the median follow-up of the cohort. We then performed a random split of the data (80%, 10%, 10%).

### Labels – Organ

For the pre-training on organ prediction, we have pooled GTEx and TCGA data ^8,26^. GTEx concerned the analysis of normal organs and TCGA the analysis of primary tumors classified by organs of origin. A random split (90%, 10%) was performed on the organ data set (no test cohort was required as the organ data were used for the pre-training task).

### Input – RNA sequencing

Inputs used to feed DeepOS were gene expression values estimated from RNA-seq. RNA-seq was the most frequent analysis commonly performed in both GTEx and TCGA and allowed to gather a maximum of examples matched with the labels described above. RNA-seq is a multistep process. RNA is first extracted from the tissue sample and sequenced. For TCGA, a vast majority of primary tumor samples came from surgical interventions while for GTEx, it came from non-diseased tissue samples from human donors. Gene expression is then estimated by the number of RNA fragments corresponding to a genome locus from a sequenced sample. TCGA and GTEx gene expressions were analyzed with the same bioinformatic pipeline from raw sequencing data and available in Recount2^21^. Gene expression was estimated in TPM (transcripts per million) with the Rail-RNA pipeline ^39^. TPM followed a Poisson distribution, so we log-transformed and scaled the data matrix using natural logarithm.

### Input – Dimension reduction on prior-knowledge

RNA-seq gene expression data is usually highly dimensional (∼23k protein coding genes plus non-coding regions) which can be a source of overfitting during the learning step of deep neural networks ^24^. To reduce the dimensions of input data, we selected important cancer-related genes based on prior-knowledge. MSigDB database ^19^ provided gene lists related to cancer hallmark and LM22 provided immune cell line specific gene lists ^20^ that are important mechanisms for cancer evolution. These two sources comprised a total of 4,499 genes also found in GTEx and TCGA RNA-seq data.

### Input – study of the gene selection

For comparison, we trained models with random selections of 4,499 input genes, excluding the ones found from cancer hallmarks and LM22. We trained those models on the same RNA-seq data, using the same workflow (hyper-parameter search and selection of the best model on validation C-index). We replicated the experiment twice, each time with different selections of random genes (#1 and #2).

### Models’ architecture

DeepOS model is a multilayers perceptron (MLP), which consists of at least three types of layers: the input layer, hidden layers and the output layer. Except for the input data, each unit uses a linear function using parameters W and b, activated by a nonlinear function such as ReLU used here for the hidden layers. Training was supervised using the backpropagation of the gradients of the error to improve model predictions, step by step, by correcting the parameters W and b. The last layer of our model was composed of linear functions.

### Loss – Survival loss

Patient survival in TCGA was calculated by the number of days to death (the event of interest) since the date of sampling. Censored patients were patients that were still alive (have not presented the event of interest) at the time of end of follow-up. Patients with good outcomes are thus more prone to be censored. Removing censored patients would influence the model to be over-pessimistic and would decrease the number of examples for training. Keeping censored patients leads to challenges in the design of a loss function to minimize. We have developed and implemented an approach to train deep learning models on survival data. With this approach, follow up was divided into a vector of B time-bins (or time intervals). In the raw data, each day of the follow up was associated with one value: 1 if the patient is alive, 0 if he is deceased and -1 if he is censored. The value of a bin was the mean of the values of each day included in this bin. The bin value ranged from -1 to 1.

For example, the bin values corresponding to a time interval of 5 days for a patient deceased at day 4 are the following:

In days: [1, 1, 1, 0, 0, 0, 0, 0, 0, 0] ⇒ bin values: [0.6, 0]

Concerning a patient censored at day 3 the bin values are:

In days: [1, 1, -1, -1, -1, -1, -1, -1, -1, -1] ⇒ bin values: [-0.2, -1]

We used the MSE, a classical loss function used to backpropagate the error of deep learning models (examples in Supplementary Table 6). The MSE is given by:

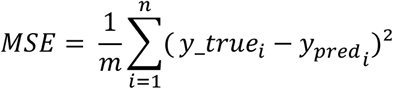

with m the number of non-censored patients and n the number of patients.

Consequently, the model output layer was designed as a vector of survival probabilities over time with the number of neurons corresponding to the number of bins. Censored values were ignored in the computation of MSE and doing so, the model was trained only on the observed follow up. The cutoff probability value for the model to predict time to death was set to 0.5 and first bin with value less than 0.5 was considered.

### Loss – Cox loss

Most of the previous studies predicting survival from RNA-seq used Cox proportional hazard model to handle censored survival data. As a control, we trained a model with DeepSurv, a MLP with a Cox log-likelihood loss function ^22^.

### Loss – For organ prediction

Categorical cross entropy was used for the organ prediction task which consisted in 38 classes.

### Training – Penalization and learning

Penalization comprises a set of classical methods to prevent overfitting during training, such as L1 and L2 regularization and dropout. Another method proposed to limit overfitting consists in adding Gaussian noise to the input data for each epoch during the training step^40^. Adam optimizer and batch normalization were also used to accelerate convergence ^41^.

### Training – Hyper-parameters optimization

Hyper-parameters are parameters controlling the MLP architecture, learning strategies and/or penalization of the learning. We have optimized the following hyper-parameters:

- The number of layers in the MLP;
- The number of nodes of the first hidden layer;
- The decrease rate of the number of unit per layer (rate by which the number of nodes of the previous layer is multiplied to determine the number of nodes of the current layer);
- The learning rates lr1 (for organ prediction task) and/or lr2 (for survival prediction task);
- The regularization parameters:
  - The standard deviation of the gaussian noise added to input data;
  - The dropout rate (continuous values within [0, 0.8]);
  - Lambda values for L1 and L2 normalization;
- The batch size;
- The number of epochs of learning.

Considering the two training tasks (organ and survival), the hyper-parameters search space had 24 dimensions. We used the Tree-structured Parzen Estimator (TPE) algorithm to train DeepOS hyper-parameters ^23^. TPE is a Bayesian approach that outperformed the traditional grid search and random search on hyper-parameters search. For each new set of hyper-parameters a new random model was fully trained. Performance metrics were calculated on the validation set(s) (for organ and/or survival). New hyper-parameters were inferred from the validation performance by the TPE algorithm. We performed 500 trials for hyper-parameters search, based on previous studies ^42^. The model with the best performance on the validation set was finally evaluated on the test set.

#### Transfer learning strategy

The transfer learning strategy for DeepOS was composed of pre-training on organ prediction and fine-tuning on survival prediction, each of these steps with independent hyper-parameters search. We used validation accuracy to select the best model on organ prediction. We then added new layers (number defined by the hyper-parameter search) and an output layer to this model. We froze the organ layers for the first fine-tuning step on survival (including hyper-parameter search), considering it as a low abstraction representation of gene expression. A second fine-tuning step (including hyper-parameter search) was performed on the same MLP with all layers unfrozen. The final survival model selection was based on the validation cohort.

#### Evaluation of the model - Metric for organ prediction

To evaluate the performance of the model on the organ task, we used classification metrics: accuracy, precision and F1 score.

#### Evaluation of the model - Metric for survival prediction

We used the concordance correlation coefficient (concordance index, or C-index) to evaluate survival models with censored data ^15,16^. C-index represents the proportion of concordant pairs divided by the total number of possible evaluation pairs. For example, if a patient A has deceased at time tA and a patient B has been censored at time tB, they can still be compared if tA<tB. If the model gives a prediction pA for patient A and pB for patient B, the pair can be qualified as concordant if pA< pB and non-concordant otherwise. If tA>tB then it is not possible to evaluate this pair and it will not count as a possible evaluation pair.

We also computed the survival AUROC using sklearn (sksurv.metrics.cumulative_dynamic_auc), which is a cumulative area under the ROC curve adapted to censored data ^43^. Finally, we used the p-value of the log-rank test to compare the predicted Kaplan-Meier survival curve to the ground truth. The log-rank test determines if two survival curves are statistically equivalent (null hypothesis) with a chi2 test. The p-value gives indication on whether we should reject the null hypothesis: the smaller it is the more two survival curves are different. Conversely, neural networks trained with Cox loss predict a risk and are barely used to predict individual survival in time; therefore log rank has not been used to date in this setting, to our knowledge.

#### Evaluation of the model - Performances by survival time

To further evaluate DeepOS predictions, we have assessed the performance depending on survival time. We have sorted the test cohort by OS and divided the cohort into 10 subgroups, each group composed of 50 patients. We have then computed the C-index of each subgroup.

#### Evaluation of the model - Learning curves

In order to evaluate the effect of the training set size on the model performances for survival prediction, we have generated learning curves. We used the validation cohort of 500 patients, given by the data split described previously. For the training set, data were iteratively and randomly added, from 500 to 5,529 samples, with steps of 500 samples. Every time a new training and hyper-parameters search was launched. The C-indexes were computed for the training and validation sets with plots for the median, the first and the third quartile.

#### Evaluation of the model – comparison to DeepSurv model

A DeepSurv model was trained, validated and tested on the same survival data split and its performances were compared to DeepOS ^22^. We used the same hyper-parameter search strategy based on the TPE algorithm. We did not perform pre-training on organ prediction with DeepSurv because of incompatibility with the Cox-loss, and because the objective was to compare our model to the existing literature.

#### Model interpretability

While still an active research field, some techniques allow interpreting MLP training. SHAP values were used, a model agnostic technique that quantifies the influence of each input on the model’s predictions ^27^. SHAP values give an input-output correlation mixed with feature importance.

#### Code and libraries

To load and process the GTEx and TCGA data we have used the R package recount2 ^21^. We have used python 3 with Keras 2.2.5 and Tensorflow 1.14, to build and train the model. Hyper-parameter search with Tree-structured Parzen Estimator (TPE) was performed with the Optuna library ^44^.

#### Code, model and data availability

The code to load and preprocess the data, together with the code to build, train and test the model is publically available on www.github.com/DITEP/DeepOS. The preprocessed data, ready to be inputted in the model, is also publically available for maximum transparency. We provided Jupiter notebooks to navigate intuitively through the steps of the analysis with results and figures included. The user that would want to run the analysis may have slightly different results as few steps are randomized (weight initialization and hyper-parameter search for example). DeepOS model trained and presented in this paper is also provided under Keras hd5 format.

## Data Availability

The code to load and preprocess the data, together with the code to build, train and test the model is publically available on www.github.com/DITEP/DeepOS. The preprocessed data, ready to be inputted in the model, is also publically available for maximum transparency.

https://jhubiostatistics.shinyapps.io/recount/

https://www.github.com/DITEP/DeepOS

## Acknowledgments

The authors are grateful to their colleagues and collaborators for their advices and support for this work and specifically: Rebecca Clodion, Roger Sun, Eric Angevin, Antoine Hollebecque, Daniel Gautheret, Stefan Michiels, Fabrice André, Andrei Zinovyev, Laurence Calzone, Emmanuel Barillot, Eric Deutsch, Jean-Yves Blay, Jean-Charles Soria, and Christophe Massard. The results shown here are based upon data generated by the TCGA Research Network: https://www.cancer.gov/tcga.

## Author contributions

Conception and design: MP, LR, HV, LV

Development of methodology: MP, LR, LV

Acquisition, analysis and/or interpretation: MP, LR, DM, LV

Writing, review and/or revision of the manuscript: all authors Supervision: LV

## Previous presentation

An intermediate version of this work has been presented at ESMO 2019 congress under the reference: Abstract 165P - Enhanced performance of prognostic estimation from TCGA RNAseq data using transfer learning. H Vanacker, E Angevin, A Hollebecque, R Sun, E Deutsch, A Zinovyev, L Calzone, E Barillot, C Massard, L Verlingue. Annals of Oncology, Volume 30, Issue Supplement_5, October 2019

## Supplementary material

### Supplementary Figures

**Supplementary Figure 1:**
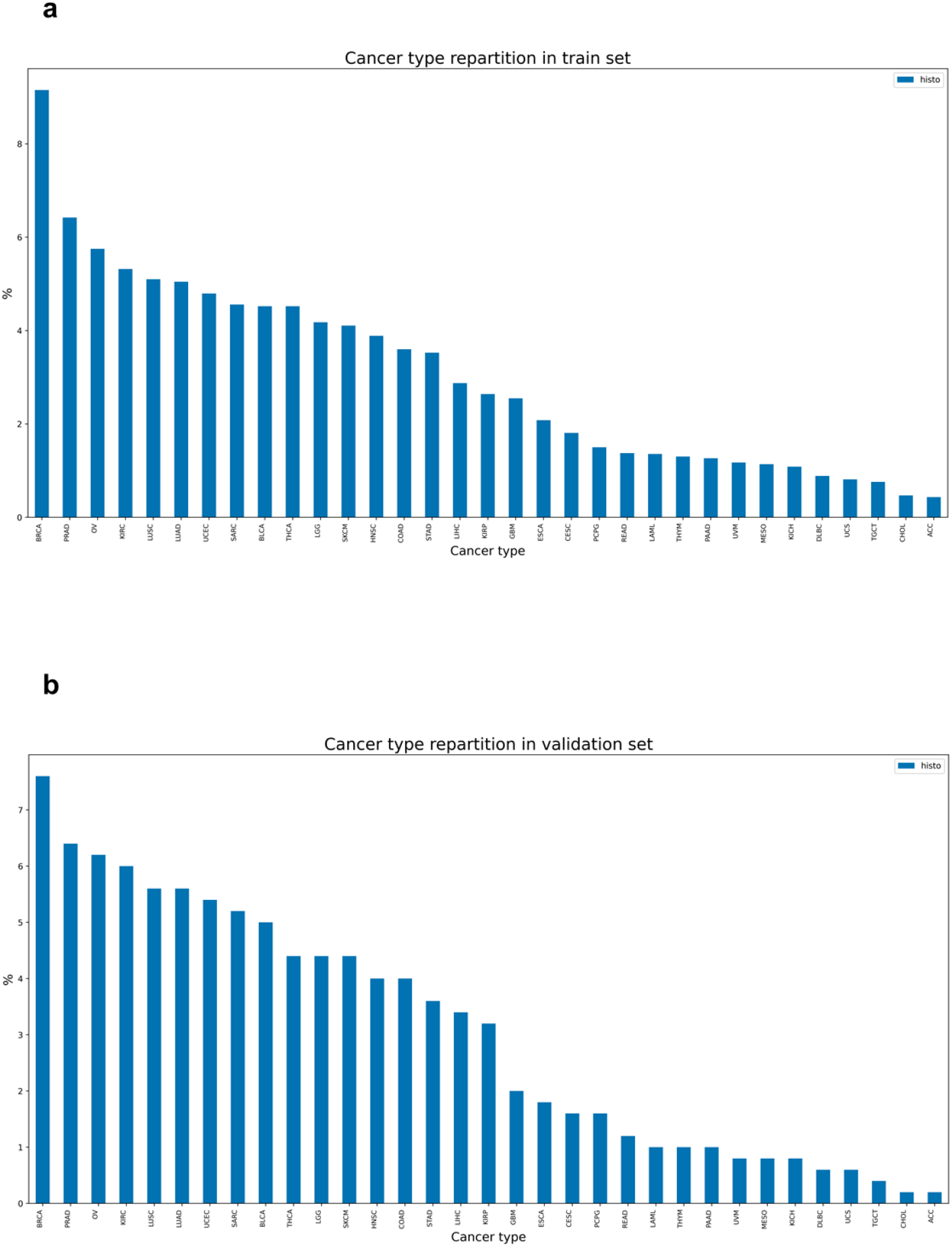
cancer type repartition in the split datasets. **a)** cancer type repartition in the train (a) and validation (b) datasets are similar. See supplementary table 3 for translation between TCGA nomenclature and histological types.

**Supplementary Figure 2:**
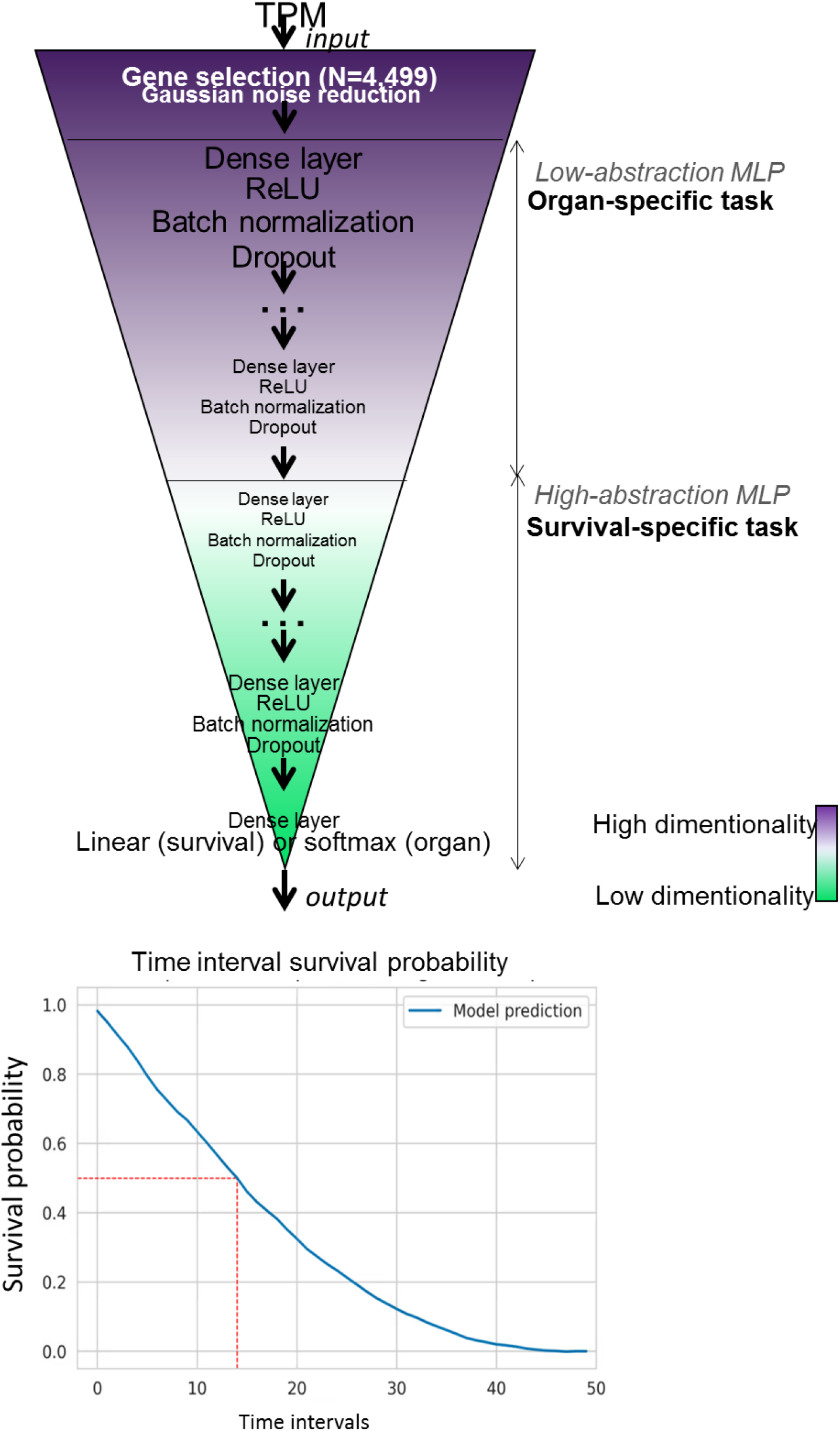
DeepOS model architecture. DeepOS is multilayer perceptron (MLP) composed of two sub-models. The first, called “Low abstraction MLP”, is specifically trained on the organ data, whereas the second, “High abstraction MLP”, is more cancer survival specific. To each layer, a ReLU function is applied, and regularization is performed (by batch normalization, dropout and L1N and L2N). DeepOS pipeline can be broken down into three sub-steps : first, the organ specific layer is generated and trained on organ data with a softmax output, then this layer is frozen and the survival specific sub-model is generated and trained on survival data, and finally, both model are unfrozen and the entire DeepOS model is trained on survival data. Survival data is transformed into time interval probabilities to be used in a mean squared error loss function to train the neural network.

**Supplementary Figure 3:**
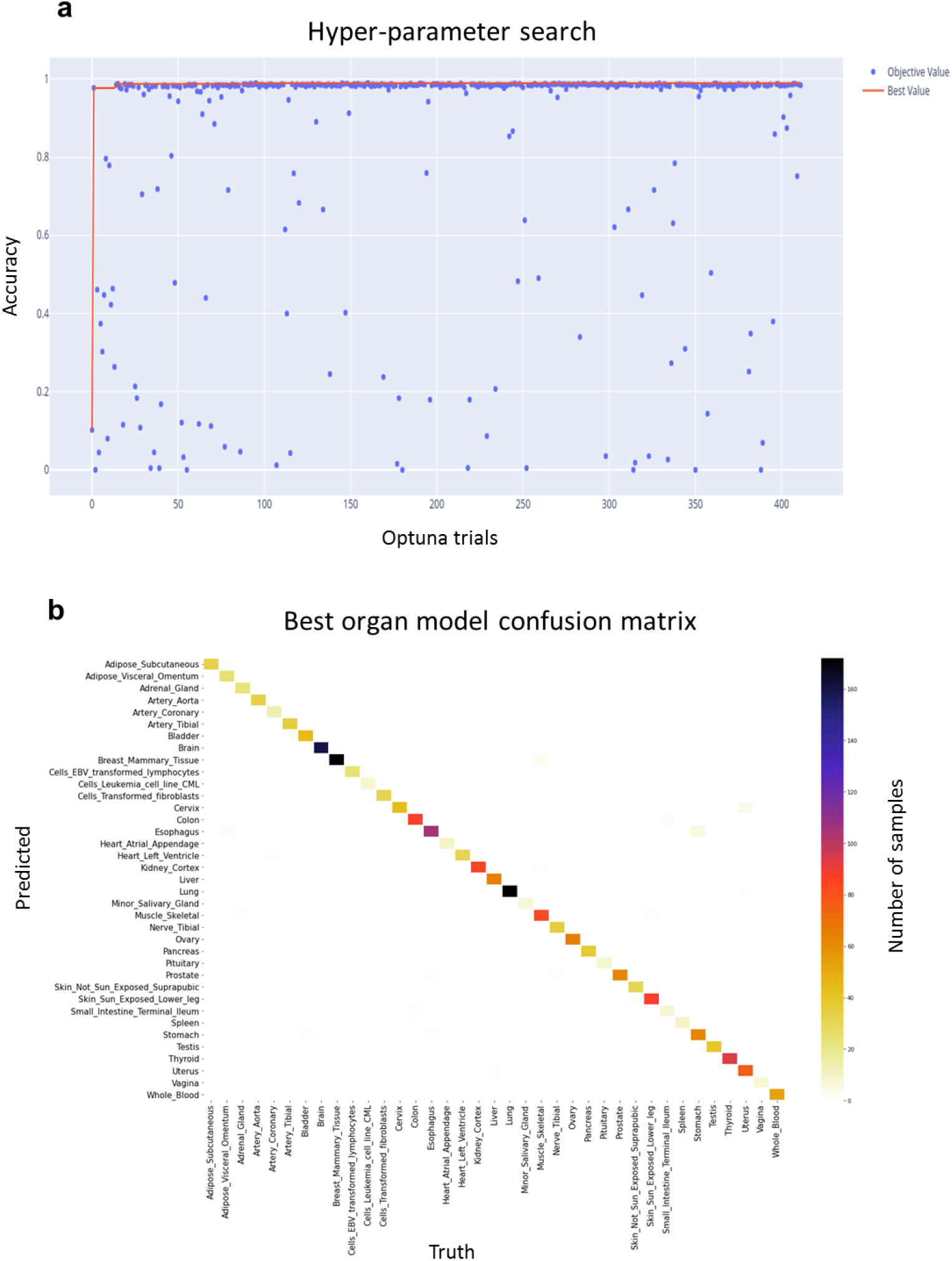
Results of the organ of origin prediction task: **a**, accuracy on the validation set for all the trials run during the hyper-parameter search procedure showing a high performance of the models overall (mean hyper-parameter search accuracy = 0.849 and standard deviation = 0.292) and **b**, confusion matrix of the best validation model predictions per organ of origin (accuracy of 0.9835, precision of 0.9842, recall of 0.9835 and F1-score of 0.9836).

**Supplementary Figure 4:**
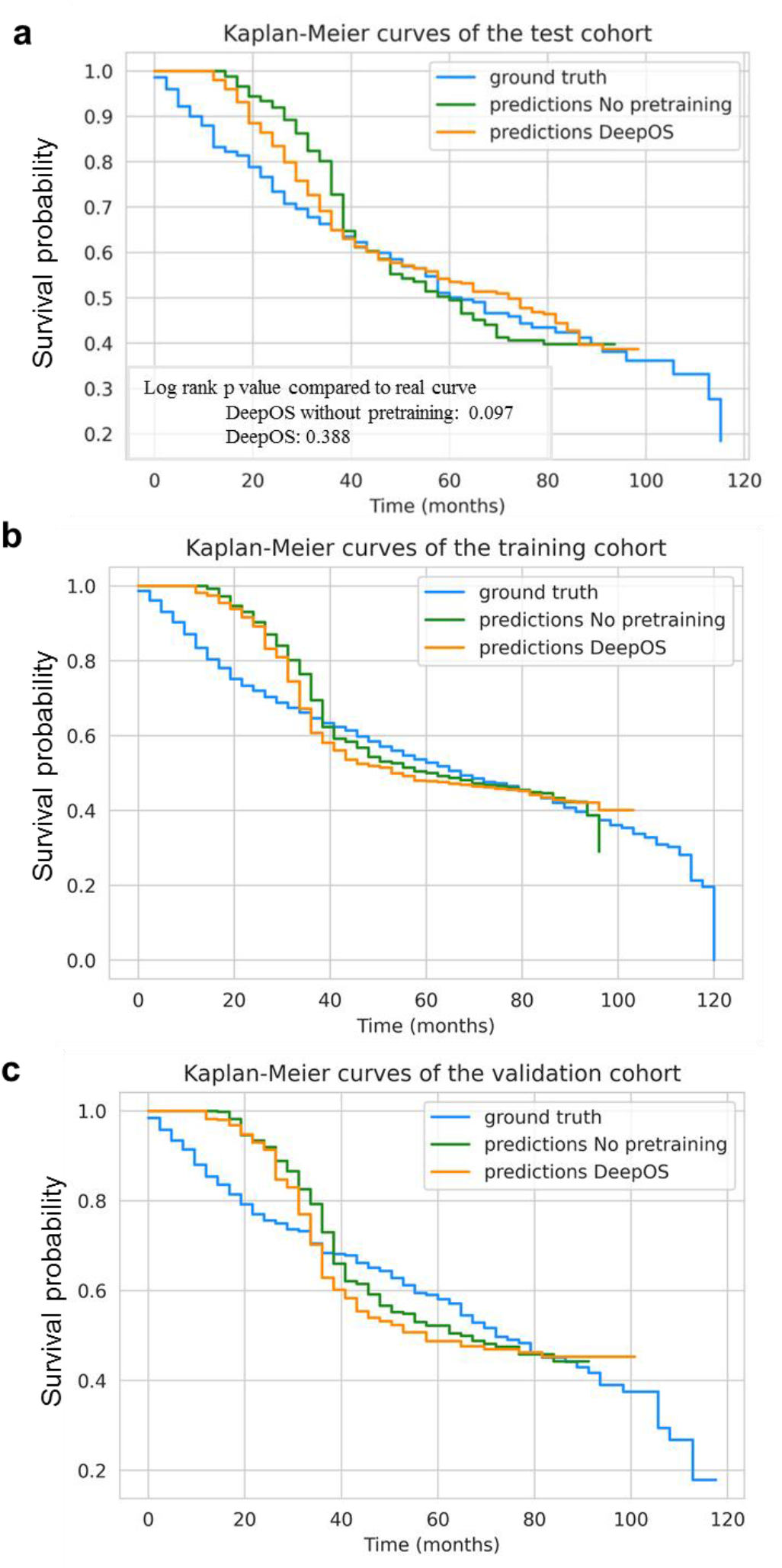
**a-c**, Kaplan-Meier survival curves predicted by DeepOS with (orange) and without (green) pre-training, compared to the observed (blue) curve of OS among patients in the test cohort (**a**), the training cohort (**b**) and the validation cohort (**c**).

**Supplementary Figure 5:**
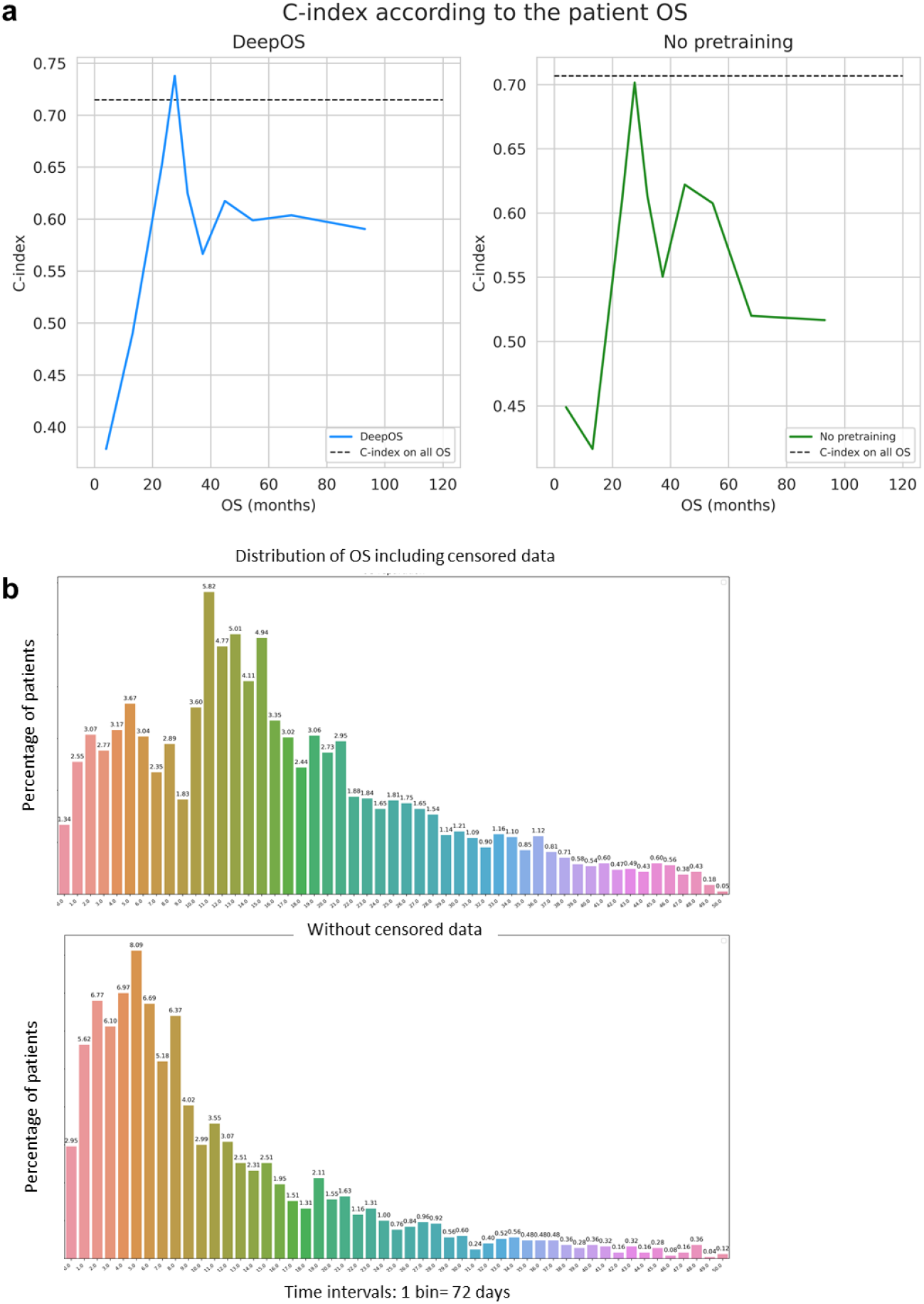
**a**, Line chart of the C-indexes computed according to the mean OS of 10 subgroups of 50 patients derived from the test cohort and predicted using DeepOS with (blue, left) and without (green, right) pre-training on organ prediction. **b**, Bar charts representing the distributions of the percentage of patients experiencing the survival event per time interval/bin, including censored events (top chart) or not (bottom chart). One time interval accounts for 72 days. OS: overall survival.

**Supplementary Figure 6:**
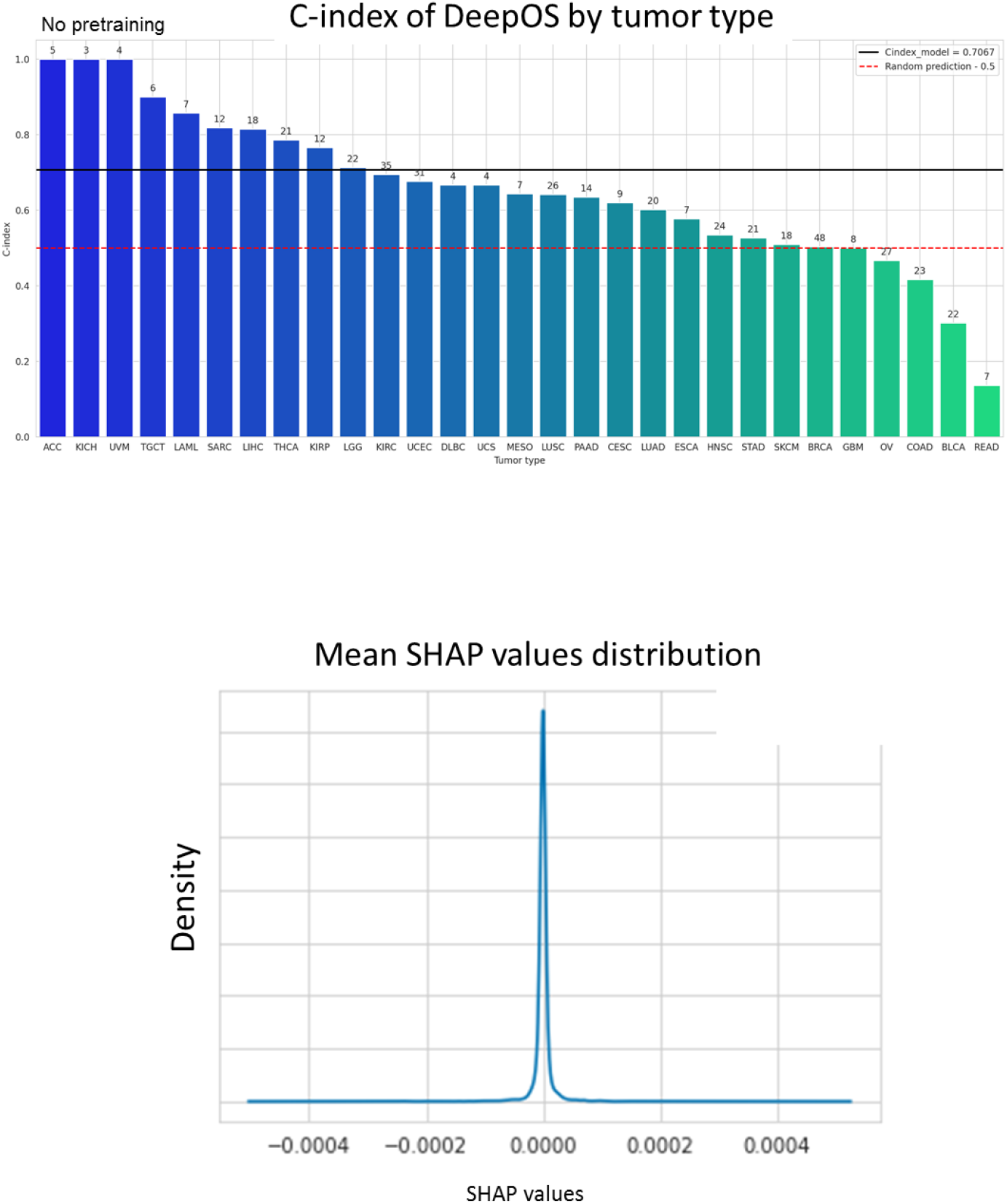
**a**, Bar chart of the C-indexes of DeepOS without pre-training according to the tumor type (compared to Figure 6a corresponding to fine-tuned DeepOS results). We considered only tumor types represented within the test cohort by at least three samples associated with uncensored survival outcome. The red dotted line indicates a C-index of 0.50 (random prediction). The black line indicates a C-index of 0.715, which refers to the median C-index of DeepOS pan-cancer predictions. Patient numbers are represented above the bars. **b**, Gaussian distribution of the mean SHAP values. We have further presented the genes with abslute mean SHAP values >0.002.

**Supplementary Figure 7:**
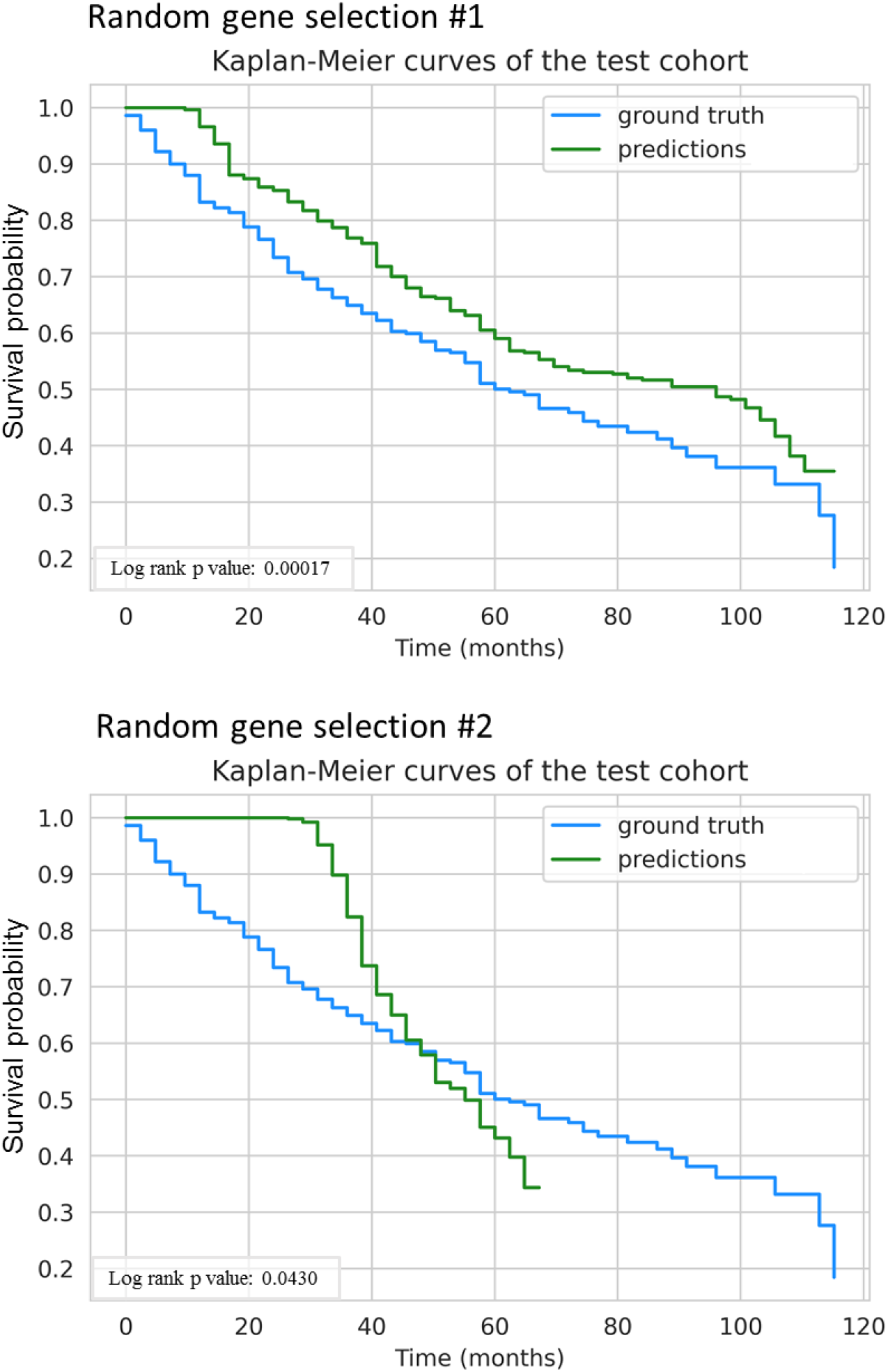
Kaplan-Meier survival curves of the test cohort predicted by the models (green) or on observed data (blue) after random selections of 4,499 genes and not used by DeepOS. Random gene selections are detailed in Supplementary Table 2. Log-rank p-values are depicted.

**Supplementary Figure 8:**
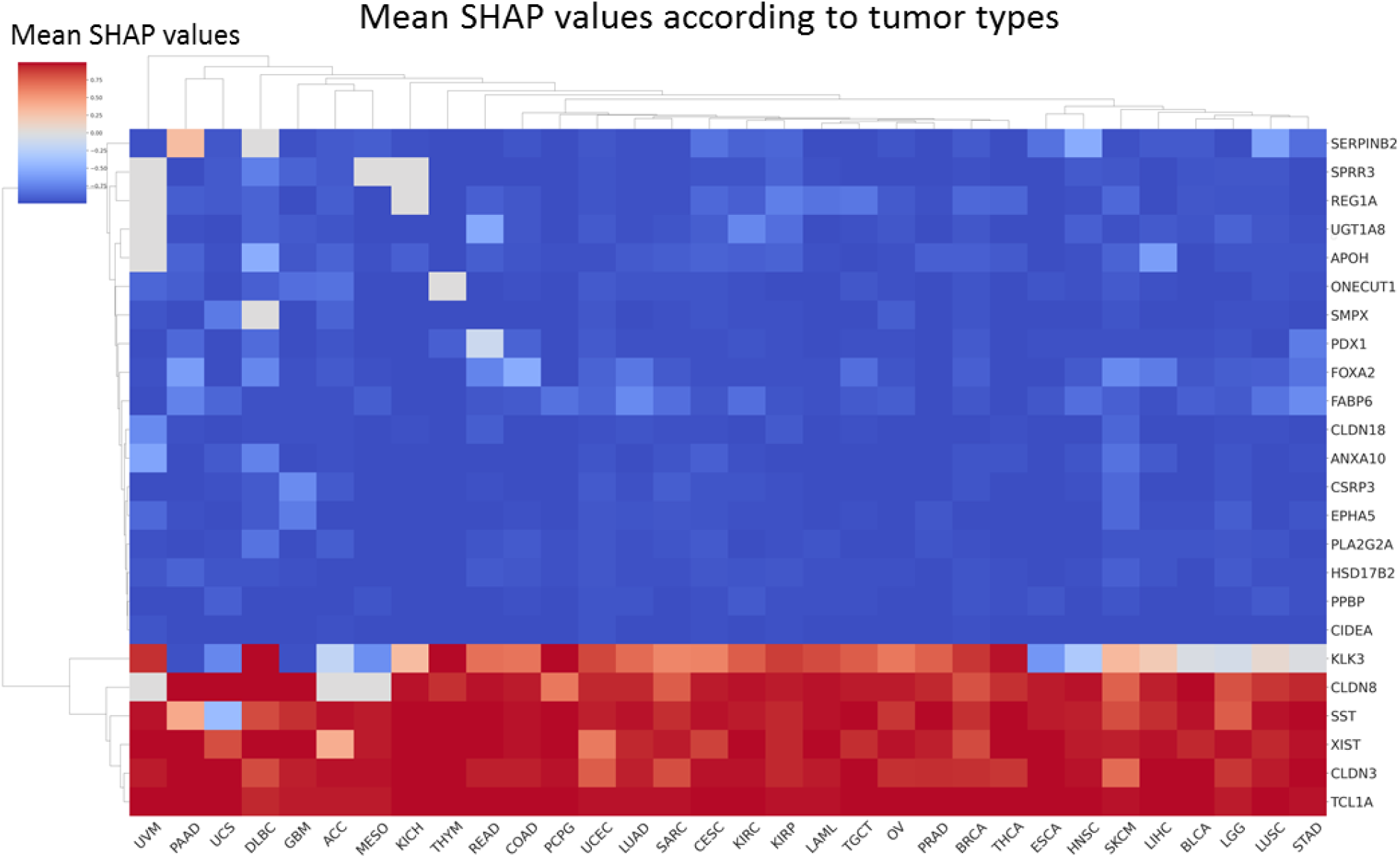
Heatmap of the mean SHAP values of the 18 genes (with absolute mean SHAP value > 0.002), according to the tumor type. Blue and red respectively indicate a positive and a negative correlation between high gene expression and poor prognosis estimation.

### Supplementary Tables

**Supplementary Table 1:**
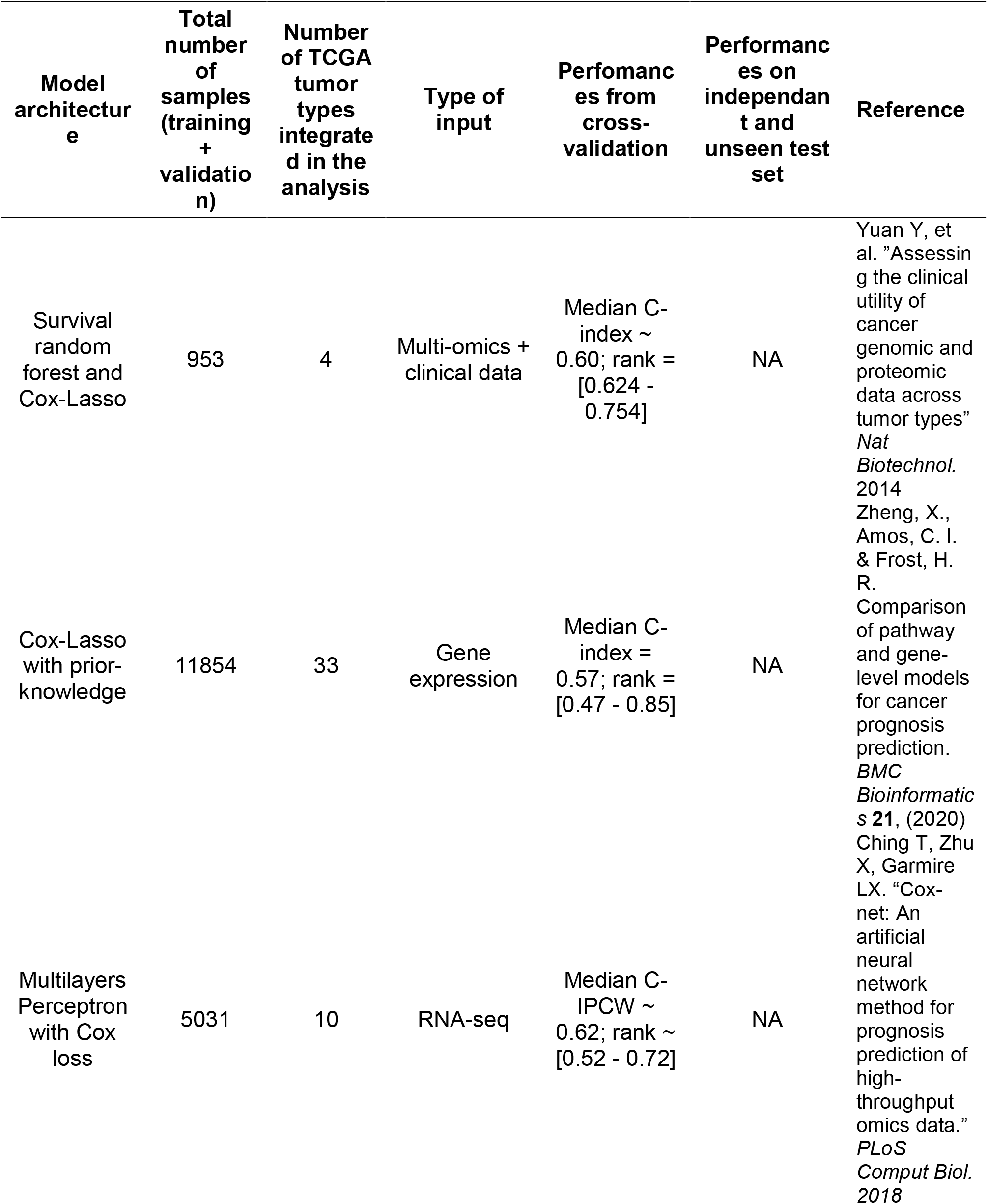
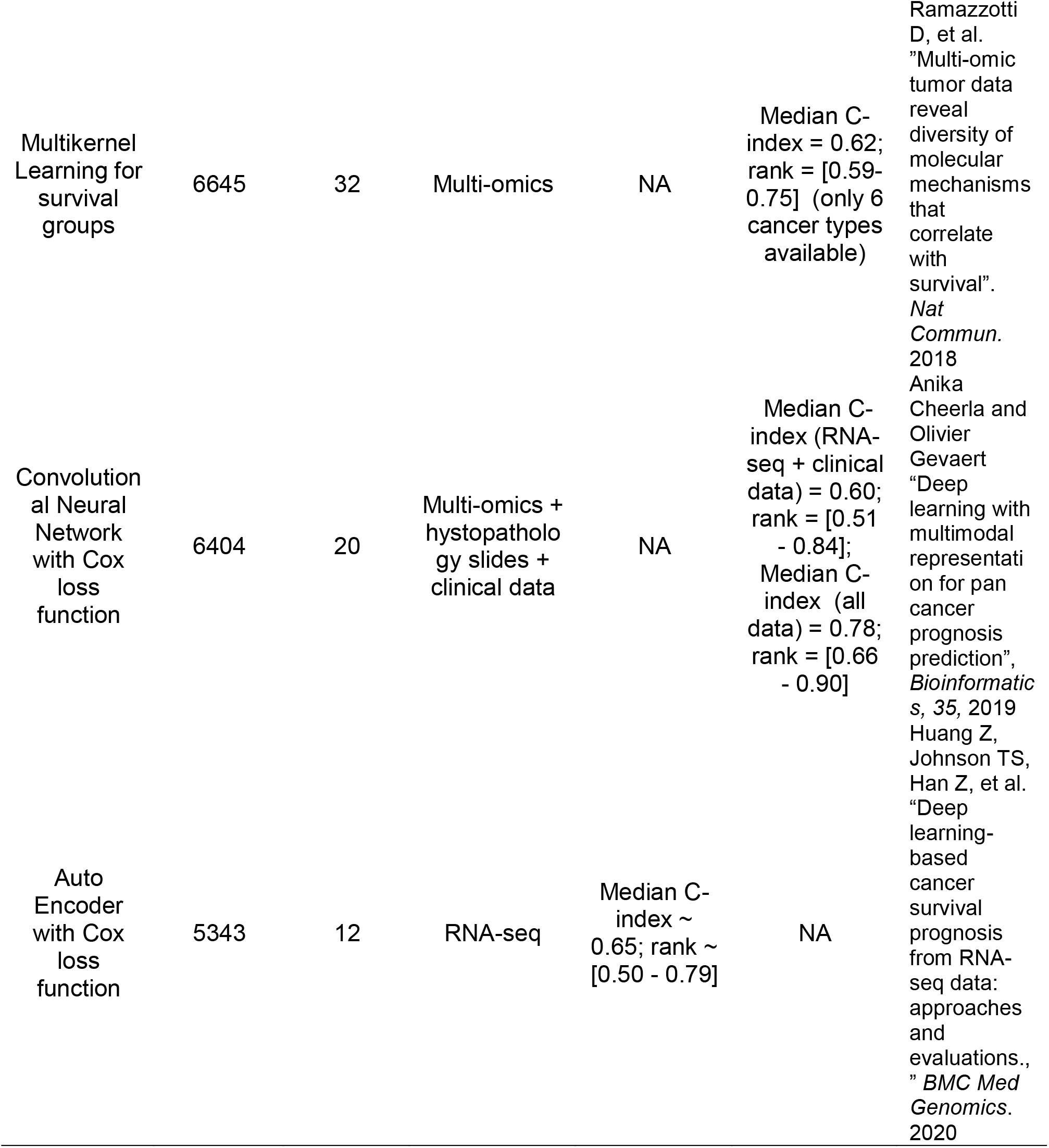
Summary of the performances of previously published Deep Learning models that used pan-cancer RNA-seq expression data from the TCGA project to predict overall survival of patients affected with cancer. NA: not available; “=” was used when the exact number was specified or could be calculated, whereas “∼” was used when the exact number was not specified and was extrapolated from a figure [too large]

**Supplementary Table 2:** Full list of the 4,499 genes related to cancer and selected to reduce dimensions in DeepOS. The indication of whether each gene was found in the MSigDB or/and LM22 database is specified by “X”. Random selections #1 and #2 depict the full gene lists that we used to study the added value of our primary gene selection.

**Supplementary Table 3:** Study abbreviation meanings of the different TCGA cohorts that were used, with related median OS (in months) and 95% confidence intervals.

| Study Abbreviation (TCGA) | Full Study Name | Median OS | 95%CI OS |
| --- | --- | --- | --- |
| ACC | Adrenocortical carcinoma | 79.2 | [52.8;inf] |
| BLCA | Bladder Urothelial Carcinoma | 24.0 | [19.2;31.2] |
| BRCA | Breast invasive carcinoma | 112.8 | [103.2;115.2] |
| CESC | Cervical squamous cell carcinoma and endocervical adenocarcinoma | 69.6 | [45.6;100.8] |
| CHOL | Cholangiocarcinoma | 24.0 | [14.4;64.8] |
| COAD | Colon adenocarcinoma | 67.2 | [55.2;93.6] |
| DLBC | Lymphoid Neoplasm Diffuse Large B-cell Lymphoma | 120.0 | [120.0;120.0] |
| ESCA | Esophageal carcinoma | 16.8 | [12.0;21.6] |
| GBM | Glioblastoma multiforme | 12.0 | [12.0;14.4] |
| HNSC | Head and Neck squamous cell carcinoma | 36.0 | [26.4;50.4] |
| KICH | Kidney Chromophobe | inf | [inf;inf] |
| KIRC | Kidney renal clear cell carcinoma | 81.6 | [72.0;120.] |
| KIRP | Kidney renal papillary cell carcinoma | inf | [86.4;inf] |
| LAML | Acute Myeloid Leukemia | 21.6 | [12.0;48.] |
| LGG | Brain Lower Grade Glioma | 64.8 | [52.8;81.6] |
| LIHC | Liver hepatocellular carcinoma | 45.6 | [28.8;57.6] |
| LUAD | Lung adenocarcinoma | 40.8 | [36.0;50.4] |
| LUSC | Lung squamous cell carcinoma | 38.4 | [33.6;50.4] |
| **MESO** | Mesothelioma | 16.8 | [14.4;24.] |
| **OV** | Ovarian serous cystadenocarcinoma | 43.2 | [38.4;45.6] |
| **PAAD** | Pancreatic adenocarcinoma | 16.8 | [12.0;19.2] |
| **PCPG** | Pheochromocytoma and Paraganglioma | inf | [inf;inf] |
| **PRAD** | Prostate adenocarcinoma | 117.6 | [115.2;117.6] |
| **READ** | Rectum adenocarcinoma | 52.8 | [48.0;inf] |
| **SARC** | Sarcoma | 60.0 | [48.0;76.8] |
| **SKCM** | Skin Cutaneous Melanoma | 50.4 | [48.0;62.4] |
| **STAD** | Stomach adenocarcinoma | 19.2 | [16.8;24.0] |
| **TGCT** | Testicular Germ Cell Tumors | inf | [inf;inf] |
| **THCA** | Thyroid carcinoma | inf | [inf;inf] |
| **THYM** | Thymoma | 115.2 | [96.0;115.2] |
| **UCEC** | Uterine Corpus Endometrial Carcinoma | 112.8 | [103.2;115.2] |
| **UCS** | Uterine Carcinosarcoma | 24.0 | [16.8;50.4] |
| **UVM** | Uveal Melanoma | 45.6 | [43.2;inf] |

**Supplementary Table 4:** Description of the training, validation and test datasets obtained from the 80%/10%/10% random splitting on selected pan-cancer RNA-seq samples from the TCGA.

|  | Number of patients | Proportion of censored patients | Median OS in months (using KM estimator) | 95%CI |
| --- | --- | --- | --- | --- |
| **Training set** | 5529 | 0.540 | 67.2 | [64.8; 72.0] |
| **Validation set** | 500 | 0.536 | 72.0 | [64.8; 88.8] |
| **Test set** | 500 | 0.546 | 62.4 | [55.2; 76.8] |

**Supplementary Table 5:**
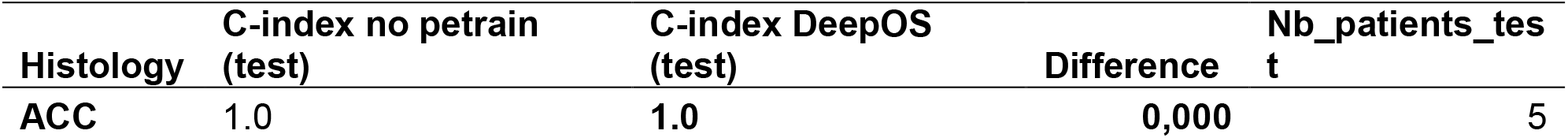
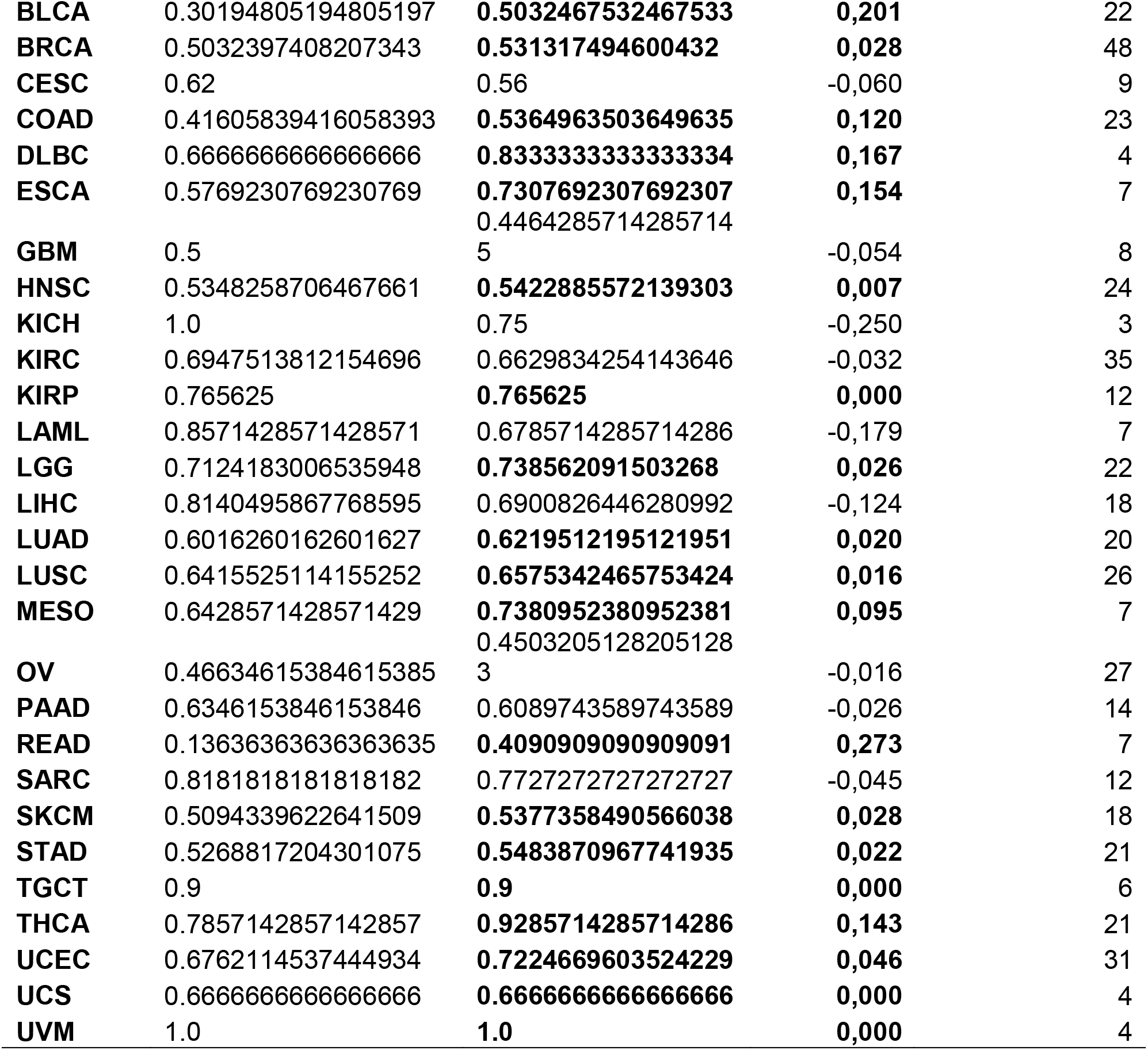
Performances (C-indexes) of DeepOS computed per tumor type on the test set, with and without pre-training.

**Supplementary Table 6:** Examples of calculation of the loss with mean squared error (MSE) for survival data. Censored values were ignored in the computation of MSE and doing so, the model was trained only on the observed follow up.

| Patient | Censored | Ground truth | Prediction | MSE |
| --- | --- | --- | --- | --- |
| 1 | No | [1,1,1,0,0] | [1.0,0.9,0.7,0.4,0.2] | 0.06 |
| 2 | Yes | [1,1,1,1,-1] | [1.0,1.0,0.9,0.8,0.7] | 0.0125 |
| 2 | Yes | [1,1,1,1,-1] | [1.0,1.0,0.9,0.8,0.1] | 0.0125 |

